# A Hybrid Pipeline for Covid-19 Screening Incorporating Lungs Segmentation and Wavelet Based Preprocessing of Chest X-Rays

**DOI:** 10.1101/2022.03.13.22272311

**Authors:** Haikal Abdulah, Benjamin Huber, Hassan Abdallah, Luigi L. Palese, Hamid Soltanian-Zadeh, Domenico L. Gatti

## Abstract

We have developed a two-module pipeline for the detection of SARS-CoV-2 from chest X-rays (CXRs). Module 1 is a traditional convnet that generates masks of the lungs overlapping the heart and large vasa. Module 2 is a hybrid convnet that preprocesses CXRs and corresponding lung masks by means of the Wavelet Scattering Transform, and passes the resulting feature maps through an Attention block and a cascade of Separable Atrous Multiscale Convolutional Residual blocks to produce a class assignment as Covid or non-Covid. Module 1 was trained on a public dataset of 6395 CXRs with radiologist annotated lung contours. Module 2 was trained on a dataset of 2362 non-Covid and 1435 Covid CXRs acquired at the Henry Ford Health System Hospital in Detroit. Six distinct cross-validation models, were combined into an ensemble model that was used to classify the CXR images of the test set. An intuitive graphic interphase allows for rapid Covid *vs*. non-Covid classification of CXRs, and generates high resolution heat maps that identify the affected lung regions.

## I. Introduction

A critical step in the fight against COVID-19 (henceforth referred to as ‘Covid’) pandemic is the screening of infected patients and the rapid recognition of those affected by Severe Acute Respiratory Syndrome CoronaVirus 2 (SARS-CoV-2). The main method used to detect Covid infection is the reverse transcriptase-polymerase chain reaction (RT-PCR). The main methods to screen for SARS-CoV-2 are chest X-ray (CXR) and computed tomography (CT) [1]. While CTs provide greater diagnostic accuracy, CXRs are more readily available and enable rapid triaging of patients. Recently, an initiative was championed by Cohen et al. [2] to provide a public repository of Covid and other respiratory distress syndrome cases with annotated chest x-ray images. Many medical centers, including ours, have developed proprietary repositories of Covid CXRs. Given the wide availability of both private and public data, a multitude of diagnostic systems based on Convolutional Neural Networks (CNNs) have been designed for automated Covid diagnosis from CXRs, in binary or multi-class assignments. It would not do justice to the countless number of reports that have been published as part of this effort to mention some and not others, and we refer the reader to some recent reviews [3-6] of the major accomplishments in this field.

Due to the still limited number of Covid CXRs available for training, a common characteristic of all proposed methods has been the adoption of transfer learning from a variety of CNNs (i.e., VGG, ResNet, DenseNet, DarkNet, SqueezeNet, Inception, Xception, each in multiple implementations of different depths) pre-trained on the collection of ImageNet (https://www.image-net.org/) data covering classes such as cars, fruits, horses, etc., but unfortunately not radiological images. Various authors have claimed that one or the other of these pre-trained CNNs gave the best results with their database of Covid images. In general, the use of CNNs with fewer layers has the advantage of lower hardware requirements, easier hyperparameters tuning, and shorter training times compared to their deeper counterparts. In fact, a survey of Covid detection performance of 15 different CNNs of 5 different architectures found that deeper pre-trained neural networks do not perform better than shallow networks [6].

Most of the published methods claim a high level of accuracy (>90%) for binary classification (Covid vs. non-Covid), and a somewhat lower accuracy (between 80% and 90%) for multiclass classification. All typically show superior or comparable performance to other reference methods considered at some point ‘state of the art’. However, some studies presented only the training accuracy (i.e., [7]), while others performed image augmentation before the train/test split (i.e., [8]), leading to some augmented versions of training images to leak in the test set. This situation suggests that one key unresolved problem in the development of effective systems for automated Covid diagnosis from CXRs is the lack of a universally accepted repository of images (as training, validation, and test sets), and the fact that most existing studies use training sets of limited-size, leading to poor generalization of their models to unseen data. A separate issue is the interpretability of the models with respect to the pertinent pathological signs in the CXR images. In fact, as impressive as the results of many studies have been in terms of classification statistics, the resulting saliency maps (usually calculated as Gradient Weighted Class Activation Maps (Grad-CAM) [9]) have been rather disappointing with activation areas often extending over both lung and non-lung regions of the CXRs (see, for example, Figure 10 in [3], Figs. 8-9 in [10], Fig. 5 in [11]), casting doubts on whether the reported classification performance is based on the recognition of particular SARS-CoV-2 features of the lungs texture or some other information (age, body shape, bone structure, sex, race, patient positioning, radiographic projection, etc.) associated with increased susceptibility to Covid infection or with specific protocols followed with Covid patients. With respect to this point, it is worth noting that deep learning systems trained to interpret X-rays and CT scans have shown a remarkable capacity to identify their subjects as Asian, Black, or White [12]. A similar conclusion was reached in [13-16], where it was noticed that although sometimes saliency maps highlight the lung fields as important, most often also highlight regions outside the lung fields, which act as confounds. These confounds are often laterality markers that originate during the CXR acquisition, which differ in style between Covid and non-Covid datasets. In other cases the saliency maps also indicate the image edges, the position of the patient shoulder and clavicle, the diaphragm and the cardiac silhouette as important for SARS-CoV-2 detection, although these regions are not used by radiologists to assess for Covid. With some datasets, similar classification results were obtained using the original CXRs or CXRs in which the lungs regions were uniformly blackened, and the classifiers was trained only on the outer part of the image [14]. Reliance on confounds from non-lung regions of the CXRs helps explain the previously observed poor generalization performance of most Covid detection models to unseen data from outside datasets. In addition to these factors, there are often also problems with the quality itself of the saliency maps. The poor multifocal discrimination of Grad-CAMs produced in some studies is likely to originate from the fact that traditional CNN architectures produce a progressive shrinking of the spatial dimensions of the feature maps, so that either the Grad-CAM maps are calculated at low resolution in the final layers of the network, and must be re-expanded to the original image resolution, or they are calculated at higher resolution in earlier layers, which however contains information very different from the downstream layers that lead to the classification output. Altogether, it is highly unlikely that in real life clinicians are going to accept a black-box model that comes with great classification statistics, but produces uninterpretable saliency maps. On this basis, it appear that the clinical efficacy of many proposed system for automated Covid diagnosis from CXRs will remain uncertain until further studies are undertaken by experienced radiologists to interpret the high-level features extracted from these systems.

In this study we did not aim to show that our Artificial Intelligence (AI) system for automatic Covid diagnosis from CXRs is better than other competing systems based on some classification statistics. Rather, our goal is to present the strategy we have adopted to circumvent the systematic problems that may invalidate the effective use of deep learning methods in the radiological detection of SARS-CoV-2 cases. Since lung segmentations were already shown to improve Covid detection in several studies [11, 13, 15-20], we designed our network so that classification would be based solely on the lung features of CXRs and would not rely on transfer learning from network trained on non-radiological images of predefined dimensions. It was also required that Grad-CAM maps would be informative and of sufficiently high resolution, so that discrete areas of the lungs could be clearly identified as having undergone Covid associated texture changes. We further aimed to develop a system that would be *production ready*, capable of differentiating patients from previously unseen images within seconds or less using only inexpensive personal computers or hand-held devices, and with an intuitive graphic interphase displaying the saliency maps produced by the model.

Here we present CXR-Net, a two-module AI pipeline for SARS-CoV-2 detection. Module 1 is based on Res-CR-Net [21], a convolutional neural network (CNN) that departs from the popular encoder-decoder U-Net architecture [22]. The idea behind the U-Net architecture is that segmentation can be conceptually decomposed in two operations: (1) semantic content extraction in the encoder arm of the neural network (NN), and (2) progressive addition/replacement of the extracted semantic content to the original size image in the decoder arm of the NN. However, it is intuitively hard to understand why these two operations cannot proceed smoothly and progressively in a pixel-wise fashion, without the need for first shrinking and then re-expanding the concept field. In our previous work on the semantic segmentation of microscopy images [21] we have shown that Res-CR-Net offers comparable or superior performance to U-net(s) in segmentation tasks. Furthermore, since its layers contain no pooling or up-sampling operations, the spatial dimensions of the feature maps at each layer remain unchanged with respect to those of the input images and of the segmentation masks used as labels or predicted by the network. For this reason, CXR-Net Module 1 (which is derived from Res-CR-Net) is completely modular, with residual blocks that can be proliferated in a straight down linear fashion as needed, and it can process images of any size and shape without changing layers size and operations, as required by U-Net. Finally, the significantly smaller number of parameters in CXR-Net Module 1 *vs*. U-Nets reduces the risk of overfitting the training set, while achieving comparable segmentation accuracy. Module 1 was trained on datasets of antero-posterior and postero-anterior (AP/PA) CXRs with radiologist annotated lung contours to generate masks of the lungs that do not overlap the heart and large vasa.

The observation that deeper neural networks pre-trained on ImageNet images usually perform more poorly than shallow networks on Covid classification tasks [6] suggests that transfer of highly specialized learning from non-radiological to radiological images may actually be deleterious. One alternative is to train a completely new network on just the database of Covid and non-Covid CXRs, with the initial layers of the network learning the general features of the X-ray images. The other alternative is to replace the image generalization provided by traditional transfer learning with the generalization provided by a wavelet based multiresolution analysis layer [23], whose coefficients are not trained and thus are not biased/optimized for classes of non-radiological images. For this reason, Module 2 was designed as a hybrid convnet in which the initial convolutional layers with learned coefficients, or the stem of a pre-trained network with its coefficient frozen, are replaced by a layer with fixed coefficients provided by the Wavelet Scattering Transform (WST) [24, 25]. Module 2 takes as inputs the patients’ CXRs and corresponding lung masks calculated by Module 1, and produces as outputs a class assignment (Covid *vs*. non-Covid) and high resolution heat maps that identify the affected lung regions. Module 2 does not depend on previously trained models, and therefore is not constrained to predefined image dimensions before training.

CXR-Net was implemented using Keras [26] deep learning library running on top of TensorFlow 2.2 [27]. It is publicly available at https://github.com/dgattiwsu/CXR-Net. g-CXR-Net, a graphic application for both Unix and Windows platforms using CXR-Net segmentation and classification engine is publicly available at https://github.com/dgattiwsu/g-CXR-Net.

## II. CXR-Net Module 1: Lung Segmentation

### A. CXRs and lung segmentation sources

All CXRs and lung masks were derived from the V7-Darwin dataset (https://darwin.v7labs.com/v7-labs/covid-19-chest-x-ray-dataset). This dataset contains 6500 images of CXRs and CTs. 5863 images are from the Kaggle dataset (https://www.kaggle.com/paultimothymooney/chest-xray-pneumonia/data). In addition to several images of normal lungs, the dataset includes 1970 images of viral pneumonia, 2816 images of bacterial pneumonia, 17 images of Pneumocystis pneumonia, 23 images of fungal pneumonia, 2 images of Chlamydophila pneumonia, and 11 images of unidentified pneumonia. Additional 517 cases of Covid pneumonia are sourced from a collaborative effort (https://github.com/ieee8023/covid-chestxray-dataset). Lung segmentations in this dataset were performed by human annotators and include lung opacities behind the heart. The final dataset (henceforth referred to as the V7 dataset) used to train Module 1 was derived from the V7-Darwin dataset by removing all sagittal views and CT scans. It consists of 6395 AP/PA CXRs, whose corresponding lung masks include the heart and large vasa contours. The dataset was split into a training and a validation set, with 6191 and 204 image/mask pairs, respectively. The original database contained 6209 unique patients, and sample selection avoided the inclusion of CXRs from the same patient in both the training and validation set. After the split, the training set contained 6018 unique patients, and the validation set contained 191 unique patients.

### B. Image resizing and ground truth labels

In order to be compatible with the image format used locally for archiving, all CXRs from the V7 dataset and the corresponding lung masks were resized to 300×340 pixels. This sizing is similar to that used in [6] to survey the Covid detection performance of 15 different CNNs of 5 different architectures. It is usually extremely challenging to differentiate pathological lesions in medical X-ray images, and image enhancement is often used to improve the lesions visibility. In our case, CXRs were histogram equalized to minimize differences in contrast/brightness within the dataset. When only a mask of the lung region was available, a complementary mask of the non-lung regions was generated. At the end of this pre-processing step, all images and ground truth binary masks were of dimensions (300×340×1), with 2 masks (one for each class, lung and non-lung) per image.

### C. Data augmentation

Each pair of image and ground truth mask(s) was sheered (0°-15° range) or rotated at random angles (0°-90° range), shifted with a random center (0-10% range), vertically or horizontally mirrored, and randomly scaled in/out (0-10% range). The parts of the image left vacant by the transformation were filled in with reflecting padding. During training, images in a batch were not shuffled, but each image underwent a different type of augmentation as determined by the consecutive calls of a random number generator starting from a fixed initial seed. Since augmentation was carried out after the splitting of training and validation sets, the model’s performance with the validation set was not biased to identify augmentation features.

### D. Architecture

A flowchart of Module 1 architecture is shown in Fig. 1, (see also for more details Supplementary Information Fig. S1). Module 1 is a *residual* network that uses *depthwise separable atrous* convolutions with *multi-scale kernels* in the residual path [28]. As stressed in [28, 29], the rationale for using multiple kernel sizes and dilations is to extract object features at various receptive field scales. In Module 1, Convolutional Residual Block (CONV RES BLOCK, Fig. 1) are repeated in a linear path along which the dimensions of the intermediate feature maps remain identical to those of the input image and of the output mask(s). In this study we used a weighted Tanimoto loss [30] for training. Weights were derived with a contour aware scheme, by replacing a step-shaped cutoff at the edges of the mask foreground with a raised border that separates touching objects of the same or different classes [31]. The unweighted Dice coefficient [32] was used as the metric to evaluate segmentation accuracy.

**Fig. 1.**
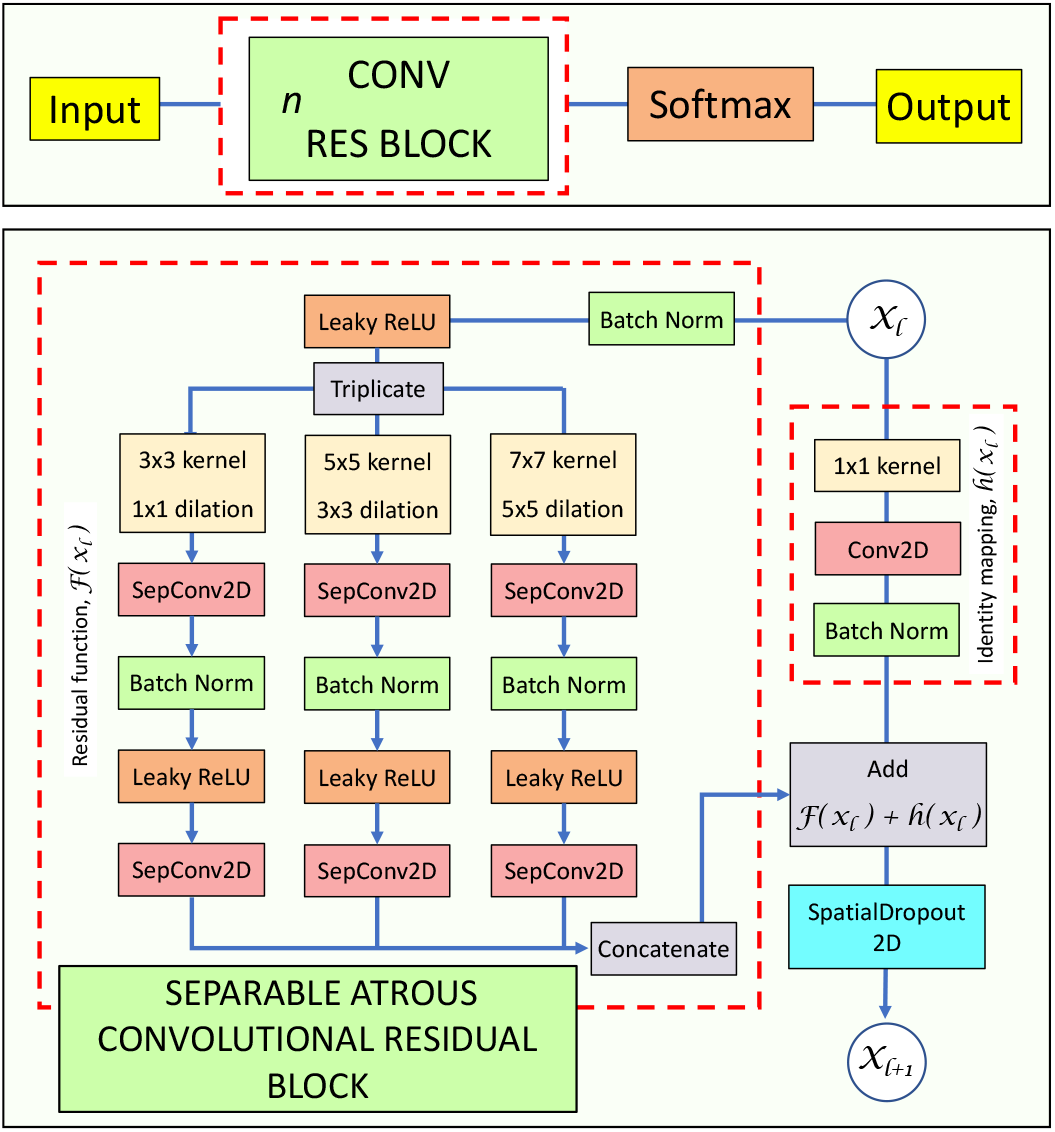
Module 1 architecture. n residual CONV RES blocks are repeated in a linear path along which the dimensions of the intermediate feature maps remain identical to those of the input image and of the output mask(s). The residual path of a CONV RES block consists of three parallel branches of separable/atrous convolutions that produce feature maps with the same spatial dimensions as the original image. Parallel branches inside the residual block are concatenated before adding them to the shortcut connection. A Spatial Dropout layer follows each residual block. In this study we have used 6 CONV RES blocks, each with kernel sizes of [3,3], [5,5], [7,7], and dilation rates of [1,1], [3,3], [5,5], respectively, with 16 filters in each residual branch, and 48 filters in the shortcut branch.

### E. Training

Module 1 was trained with the V7 dataset using the Adam optimizer [33] in its default Tensorflow parameterization. Other hyperparameters were left unchanged from those identified during the testing of Res-CR-Net with microscopy images [21], the only difference being that the final Long Short Term (LSTM) memory block of Res-CR-Net was removed from the architecture of Module 1 to decrease the training time with the large number of images in the V7 dataset. Since no hyperparameter tuning was carried out, the validation set effectively acted as a test set. Based on the trend of the validation loss and accuracy (Fig. 2), training was stopped after 100 epochs. The total number of parameters refined was 58,309. Each epoch consisted of 516 training batches of 12 images each, and 17 validation batches also of 12 images each. Run time was 2.814 s/batch (forward + backward propagation), 322 ms/batch (forward propagation only) using 2 Nvidia Tesla V100 GPUs of Wayne State University High Performance Computing (WSU-HPC) grid. Memory usage was 13.82 Gb/batch. The number of million floating point operations per second (MFLOPS) in a forward pass was 11,485.

**Fig. 2.**
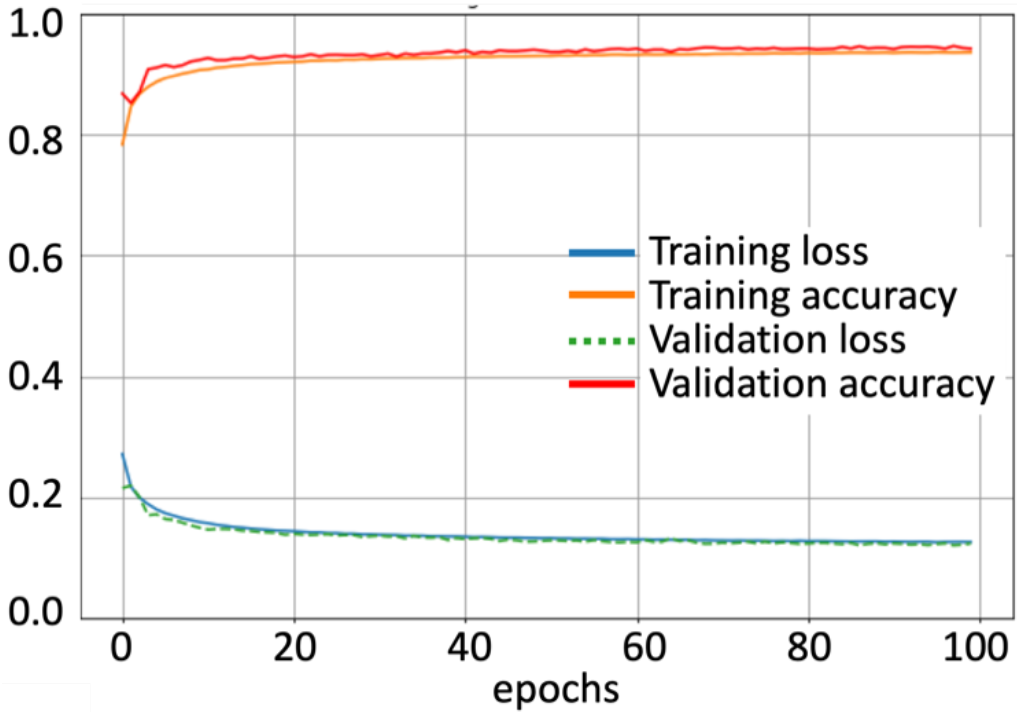
Training and validation weighted Tanimoto Loss and Accuracy (as Dice Coefficient) vs. epochs for Module 1 processing of V7 images.

In every epoch, the network trained on 6192 different augmented images, and the corresponding augmented masks as labels. Upon training, Module 1 achieved ∼94% segmentation accuracy on the images of the validation set. The training history showed no overfitting of the training vs. the validation set (Fig. 2). The segmentation task was to identify the regions occupied by the lungs with exclusion of the skeletal structures visible in the CXR, but including cardiovascular components and opacities due to underlying pathologies (Fig. 3). Module 1 performed very well with the V7 training and validation datasets, achieving a score of 0.96 with respect to each of the metrics used to evaluate the network performance. These were:

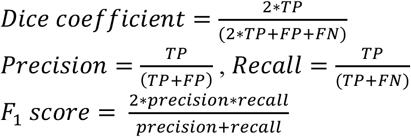

where: TP = true positive, FP = false positive, FN = false negative

**Fig. 3.**
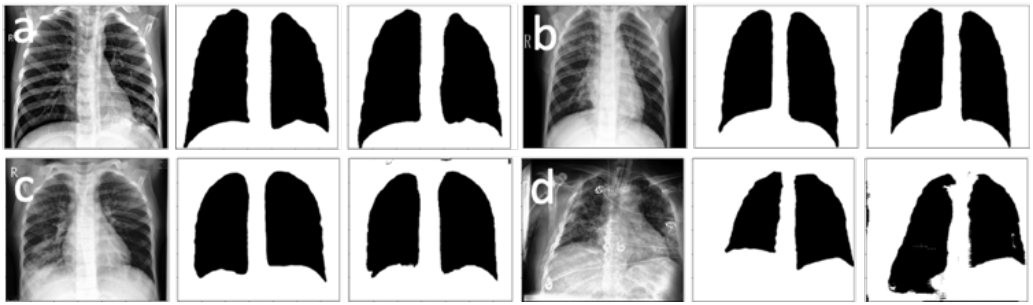
Examples of lung segmentation from the V7 validation subset. Left panels, CXR, Center panels, ground truth mask, Right panels, Module 1 predicted mask thresholded at 0.5 value. Rows a-c show three cases in which the mask is very similar to the ground truth mask. Row d shows a case in which the predicted mask includes areas of the images that belong to the abdomen.

### F. Generation of lung masks for the HFHS dataset

Module 1 trained on the V7 dataset was used to generate lung masks for the CXRs in a dataset of AP/PA CXRs from non-Covid and RT-PCR confirmed Covid patients acquired at the Henry Ford Health System (HFHS) Hospital in Detroit (the ‘HFHS dataset’). All non-Covid CXRs were from pre-Covid era (2018-2019), and included images from both normal lungs and lungs affected by non-Covid pneumonia or other lung pathologies. HFHS CXR images were resized to 300×340 pixels and histogram equalized to minimize differences in contrast/brightness within the datasets, prior to passing through CXR-Net Module 1 to calculate the corresponding masks of the lung regions. At the end of this pre-processing step, all images and masks were of dimensions (300×340×1). Example of CXRs from this dataset, and the corresponding lung masks generated by Module 1 are shown in Fig. 4. In all cases, mask values were kept in the original [0,1] floating point range representing the probability of an image region to be part of the lungs, without thresholding the mask at 0.5 value for conversion to a binary mask. Module 1 performance with the previously unseen CXRs of the HFHS dataset was very good, as judged by visual inspection of the masks, although in this case lung contours validated by radiologists were not available.

**Fig. 4.**
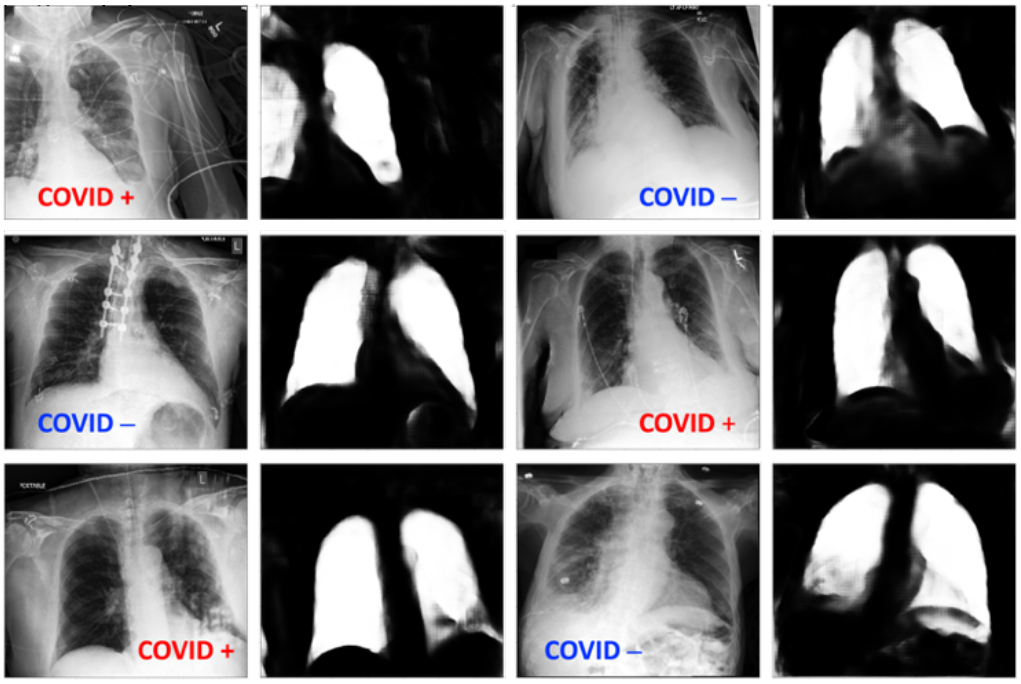
Examples of lung segmentation in CXR images from the HFHS dataset. CXRs were labeled by radiologists as Covid or non-Covid. Module 1 generated floating point lung masks are shown next to each CXR.

## III. CXR-Net Module 2: Covid vs. non-Covid classification

### A. CXRs sources

CXR-Net Module 2 was trained against 2265 CXRs from 1313 unique patients (1417 non-Covid, of which 1075 from unique patients, 848 Covid, of which 238 from unique patients), and tested against 1532 CXRs from 895 unique patients (945 non-Covid, of which 735 from unique patients, 587 Covid, of which 160 from unique patients)), all from the HFHS dataset. CXRs were split into training and test set by avoiding inclusion of CXRs from the same patient in both sets. CXRs were resized to 300×340 pixels and histogram equalized. Module 1 calculated lung masks values were in the [0,1] floating point range. Both training (2265 CXRs) and test images (1532 CXRs) were standardized to the common mean and standard deviation of the training set. The training set, was further split for 6-fold cross-validation into 6 distinct sets of 1887 training images and 378 validation images, keeping in each set the same ratio of Covid to non-Covid images of the entire set of 2265 images. During training, CXRs were further assigned weights accounting for class imbalance (Covid *vs*. non-Covid).

### B. Architecture

Module 2 is a hybrid convnet in which the first convolutional layer is a layer with fixed coefficients provided by the Wavelet Scattering Transform (WST) [24, 25]. A scattering network belongs to the class of CNNs whose filters are fixed as wavelets [34]. Thus, an important distinction between the scattering transform and a deep learning framework is that the filters are defined a priori as opposed to being learned. The construction of the scattering network relies on few parameters and is stable to a large class of geometric transformations [35], making its output a suitable generic representation of an image. A wavelet scattering framework enables the derivation from image data of low-variance features insensitive to translations of the input on a user-defined invariance scale; in the 2D case, they are also insensitive to rotations. The scattering framework uses predefined wavelet and scaling filters. Efficient algorithms for the 2D scattering transform have been implemented as a NN layer in Keras/Tensorflow *via* the Kymatio software [36].

A wavelet is an integrable and localized function in the Fourier and space domain, with zero mean. A family of wavelets is obtained by dilating a complex mother wavelet *ψ* as *ψ*_*j,θ*_where *j* is a dilation scale and *θ* a rotation. *J* and *L* are integers parameterizing the dilation scale and the angular range. A scattering transform (*J* being its spatial scale) generates features in an iterative fashion. Given a grayscale image *x*, and a local averaging filter *φ*_*J*_ with a spatial window of scale 2^*J*^, we obtain the *zeroth order* scattering coefficients of *x* as:

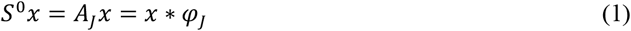

(where * indicates here convolution). This operation leads to a down-sampling of scale 2^*J*^. For example, in the case of a grayscale image of dimensions *N* × *N, S*^0^*x* is a feature map of resolution *N*/2^*J*^ × *N*/2^*J*^ with a single channel. The zeroth order scattering transform is invariant to translations smaller than 2^*J*^ (which for this reason is often referred to as the *invariance scale* of the transform), but also results in a loss of high frequencies, which are necessary to discriminate signals. However, the information is recovered when computing the coefficients in the next stage. Each stage consists of three operations:

1. *Take the wavelet transform of the input data with each wavelet filter in the filter bank.* A wavelet transform, *W*, is the convolution of a signal with a family of wavelets, with an appropriate down-sampling. The first order wavelet transform of *x* is:

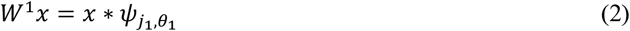
2. *Take the modulus of each of the filtered outputs*
3. *Average each of the moduli with the scaling filter φ*_*J*_.

Thus, the *first order* scattering coefficients can be calculated as:

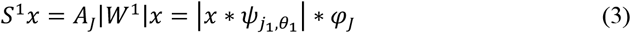

In a grayscale image (i.e., a CXR from Fig. 4), *S*^1^*x* is a feature map of resolution *N*/2^*J*^ × *N*/2^*J*^ with *JL* channels. The use of averaging also in this first order, generates invariance to translation up to 2^*J*^. To recover some of the high-frequencies lost due to the averaging applied on the first order coefficients, we apply a second wavelet transform *W*^1^ (with the same filters as *W*^1^) to each channel of the first-order scatterings, *before* the averaging step. This leads to the *second-order* scattering coefficients, calculated as:

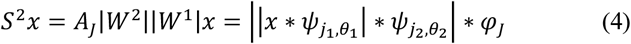

In our grayscale image example, *S*^2^*x* is a feature map of resolution *N*/2^*J*^ × *N*/2^*J*^ with *J*(*J* − 1)*L*^2^ channels. The energy of higher order scatterings rapidly converges to 0. Thus, for most applications, a framework with two wavelet filter banks is sufficient. For this reason, the final scattering coefficient *S*^*J*^*x*, which are the low-variance features derived from the image, correspond to the concatenation of the order 0, 1 and 2 scattering coefficients. *S*^*J*^*x* is a feature map with 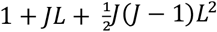 channels, down-sampled by a factor of 2^*J*^ with respect to the original image. This representation has proven to linearize small deformations of images, to be non-expansive, and almost complete [35, 37], which makes it an ideal input to a deep convolutional network. An example, with *J*=2 and *L*=6, of the wavelets for each scales and angles that would be used for an image *x* of dimensions 30 × 34 (1/10 of those used in this study) is shown in Fig. 5a. The corresponding *scattering transform* tree is shown in Fig. 5b. The WST should not be confused with the Discrete Wavelet Transform (DWT), a different wavelet based technique that was recently used in another hybrid convnet designed for Covid detection [38]. DWT is an earlier technique developed by Mallat [23] to analyze signals into progressively finer nested octave bands. Later on, the same Mallat proposed WST [35] as a more effective multiresolution image analysis technique for deep learning systems. In early attempts to construct a hybrid convnet we also used the DWT, but found that the WST offered superior performance, in agreement with Mallat’s recommendation.

**Fig. 5.**
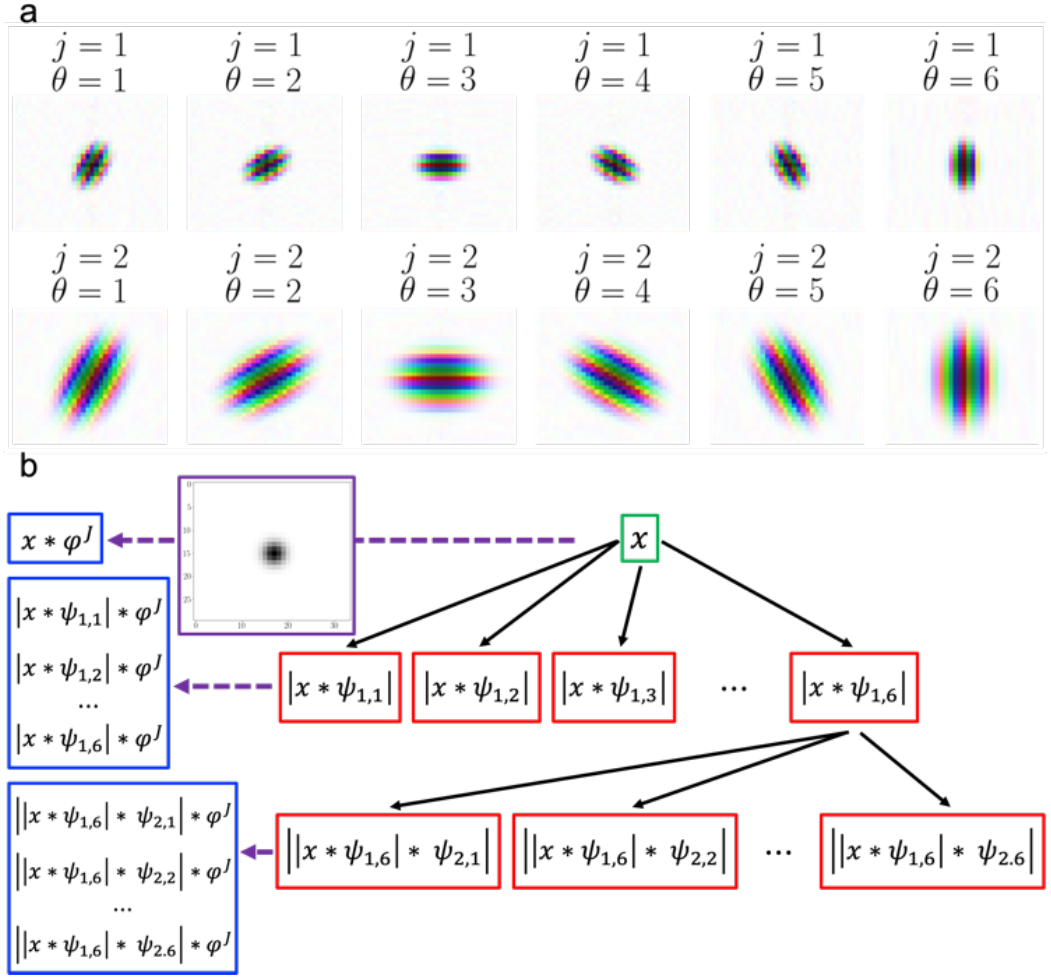
Wavelet scattering transform. Panel a: Wavelets for each scales and angles used for the transform of an image *x* of dimensions 30 × 34 with J=2 and L=6. Color saturation and color hue denote complex magnitude and complex phase, respectively. Panel b: a tree showing the calculation of the different channels of the scattering transform. Boxes with red outline represent the wavelet transform of the image with each wavelet filter and with modulus applied. The purple dashed arrows represent the averaging operation; the averaging filter is shown in the box with purple outline. The corresponding scattering coefficients for orders 0, 1, and 2 are shown in the boxes with blue outline. Additional scattering coefficients derived from the wavelet transform of the input image *x* with the 2^nd^ bank of wavelet filters are not shown.

A flowchart of the architecture of CXR-Net Module 2 in the *default* configuration of the pipeline (see Table 2, Section labeled Module 1+2) is shown in Fig. 6. Module 2 takes as inputs the patients’ CXRs and corresponding floating point lung masks (with [0,1] range representing the probability of an image region to be part of the lungs) calculated by Module 1, and produces as outputs a class assignment (Covid *vs*. non-Covid). For input images of dimensions 300 × 340, the WAVELET SCATTERING TRANSFORM (WST) block (whose coefficients are not trainable, and determined only by the scale *J* and the number of rotations *L*), with *J*=2 and *L*=6 produces two outputs with down-sampling by 2^*J*^ = 4. The 1st output is a feature map of dimensions 75 × 85 × 50. The first 49 channels are the scattering transform. The 50th channel is the down-sampled floating point mask (Fig. 7). The 2nd output is a binary mask of dimensions 75 × 85 × 1 of the lung regions derived from the corresponding floating mask by thresholding at 0.5 and down-sampling by 2^*J*^. These outputs are passed to an ATTENTION block [39, 40] that calculates a map representing a cross attention between the lung masks and the input CXR image (Supplementary Information Fig. S2). This map is concatenated to the wavelet scattering transform output from the WST block producing a feature map of dimensions 75 × 85 × 51. Upon passing through a Spatial Dropout and a Batch Normalization layer this map becomes the input to a residual CONV RES block (of the same architecture as described for Module 1) repeated 3 times in a linear path along which the dimensions of the intermediate feature maps remain identical to those of the input map. For these blocks, we have used a kernel size of [3,3] with dilation rates of [1,1], [2,2], [3,3], respectively, with 17 filters in each residual branch, and 51 filters in the shortcut branch. The final CONV RES block outputs a feature map of dimensions 75 × 85 × 2, with channels corresponding to the Covid and non-Covid classes. Both channels are first multiplied by the binary mask of the lung region (2nd input to the CONV RES blocks), then globally averaged before passing to a Softmax layer for Covid vs. non-Covid classification. The ATTENTION block requires the training of ∼1,100 parameters. The CONV RES blocks require the training of fewer than 20,000 parameters. It is worth noting that another type of *residual* network using *depthwise separable atrous* convolutions with *multi-scale kernels* in the residual path, named ‘Dilated and Depthwise separable Convolutional Neural Network (DDCNN)’ has been proposed recently by Li et al. [10] for the diagnosis of Covid-19 from CXRs. However, while DDCNN progressively reduces the spatial dimensions of the feature maps as they pass through several pooling layers, the feature maps of CXR-Net Module 2 retain the same dimensions (75 × 85 × 51) of the input image (as downsized by the WST block), with the exception of the feature map produced by the last block, which is of dimensions 75 × 85 × 2, as it is used for binary classification (Fig. 6).

**Fig. 6.**
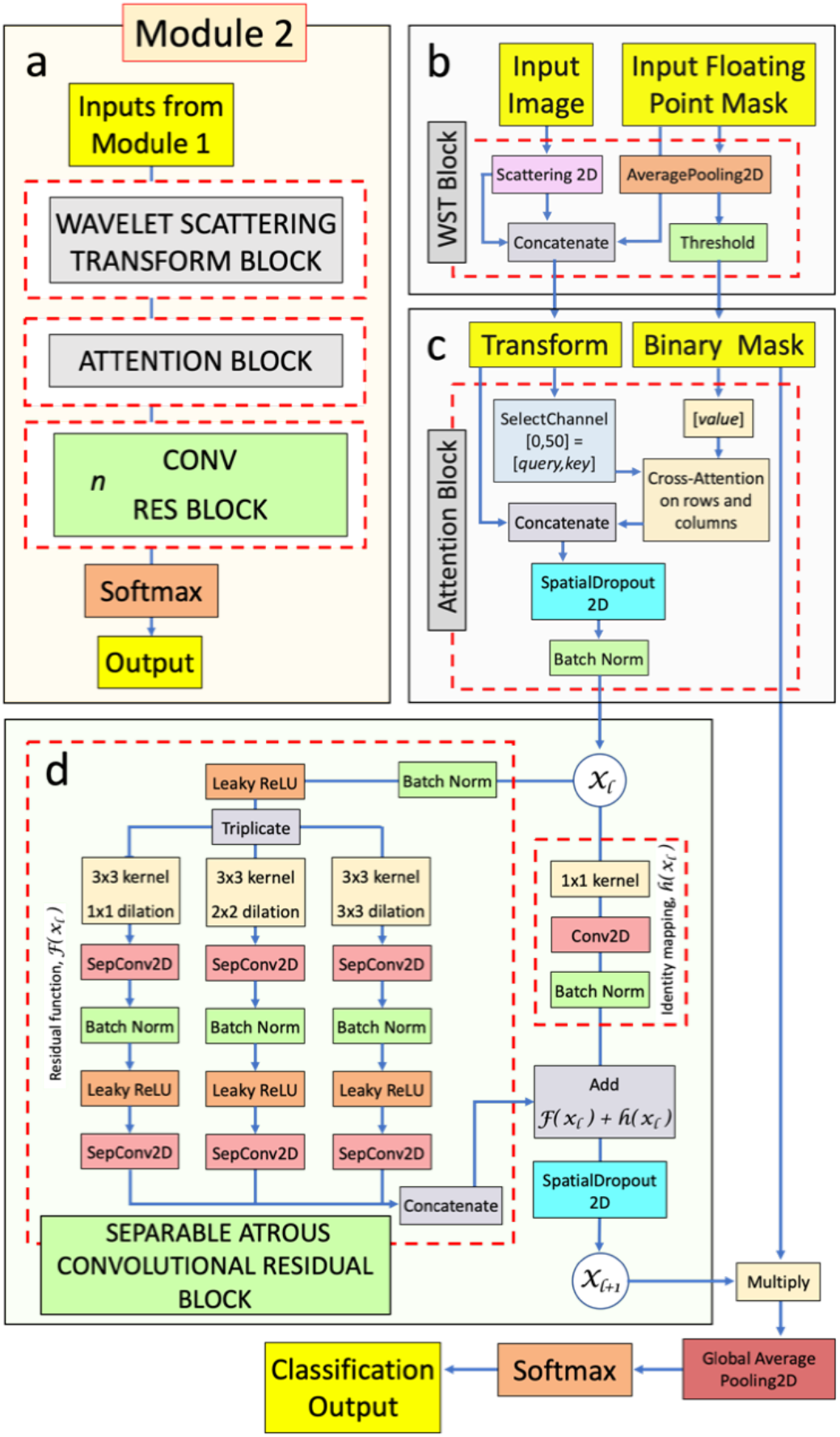
Module 2 architecture (default configuration, Table 1). Panel a: overall architecture. Panel b: details of the WST BLOCK. Panel c: details of the ATTENTION BLOCK. The box labeled ‘Cross-Attention on rows and columns’ consists of two MultiHeadAttention layers working with the *query, key*, and *value* matrices for rows attention, and their transpose for columns attention. Panel d: details of the CONV RES blocks used in this module. In this case we have used 3 CONV RES blocks, each with kernel size of [3,3], and dilation rates of [1,1], [2,2], [3,3], respectively, with 17 filters in each residual branch, and 51 filters in the shortcut branch.

**Fig. 7.**
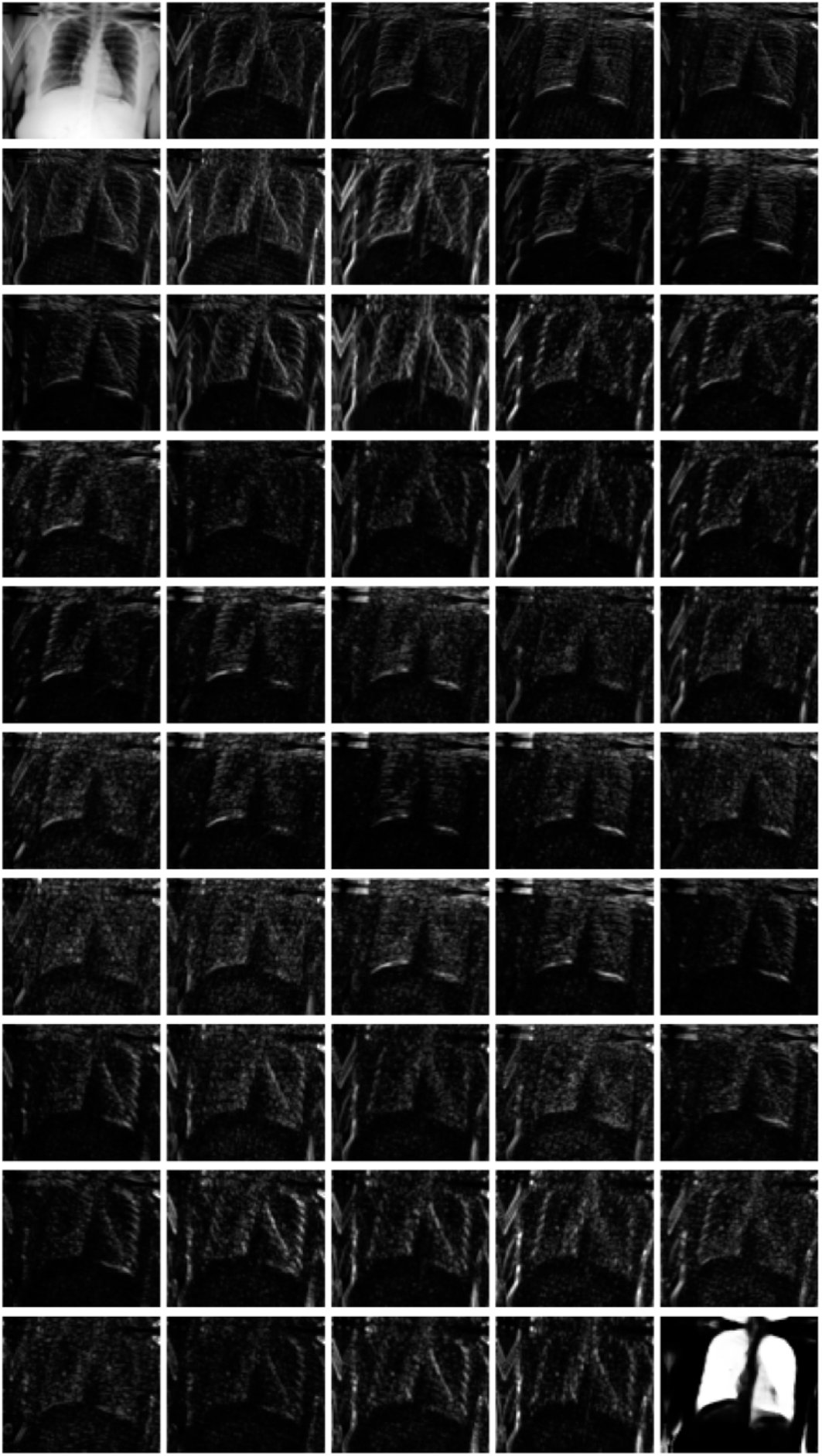
Wavelet Scattering Transform. An example of the feature map of dimensions 75 × 85 × 49 produced by the Wavelet Scattering Transform Block. The top-left panel is the down-sampled input CXR, the bottom right panel is an extra channel with the down-sampled floating point mask. All other panels represent the transform output at different scales and rotations of the analytical wavelet.

### C. Training

In separate runs, different components of CXR-Net were selectively included/excluded in order to ascertain their relative contribution to the final classification performance of the pipeline (Table 1). The following CXR-Net architecture configurations were evaluated:

1. *Default* configuration (Table 1, Section labeled ‘CONF 1’, Supplementary Information Fig. S3): Module 2 was trained with Module 1 generated lung masks. In this case Module 1 had its parameters frozen, and only the parameters of Module 2 were progressively refined. The final *Covid vs. non-Covid* classification represents the outcome from both modules acting sequentially.
2. Training as in 1., but without the ATTENTION block in Module 2 (Table 1, Section labeled ‘CONF 2’, Supplementary Information Fig. S4). The feature map channel produced by this block (Fig. 6c) was replaced with a channel containing the binary mask of the lungs.
3. Training as in 1., but with the WST block in Module 2 replaced by a CONV RES block (Table 1, Section labeled ‘CONF 3’, Supplementary Information Fig. S5).
4. Training as in 1., but with the WST block in Module 2 replaced by 2 CONV RES blocks (Table 1, Section labeled ‘CONF 4’, Supplementary Information Fig. S6).
5. Module 2 was trained without lung masks calculated by Module 1 (Table 1, Section labeled ‘CONF 5, Supplementary Information Fig. S7): in this case also the ATTENTION block was removed, as it uses the lung masks, and the channel produced by this block (Fig. 6c) was replaced with a channel with all pixels equal to 1. However, lung masks are optionally still used to calculate heat maps.

**Table 1.** Performance of Different CXR-Net Configurations. Complexity measures (parameters, mflops, memory, eval. time) are shown for Module 2 contribution only. The contribution from Module 1 (section II, *E*, above) must be added to obtain the computational complexity of the entire pipeline. ^a^The 1st memory value reported for the single model reflects the space needed for both forward and back-propagation (training) with a batch of images of the training set. The 2^nd^ memory value reflects the space needed for just forward propagation (‘evaluation’) with a batch of images of the test set. Since all the coefficients of the ensemble model are frozen, only the memory space needed for forward propagation is reported. ^b^Evaluation time refers to the time required for forward propagation with a batch of images of the test set, and with all calculations split between 2 Tesla V-100 GPU cards. In all cases, evaluation of either the validation or the test set by a single model or the ensemble model was carried out with the original non-augmented images. ^c^MFLOPS, million floating point operations per second.

| **CONF 1: Module 1 masks + Module 2** |  |  |  |
| --- | --- | --- | --- |
|  | Single Model | Ensemble model |  |
| Parameters | 20,467 | 122,802 |  |
| Memory<sup>a</sup> | 0.408 Gb/batch – 20.14 Mb/batch | 30.37 Mb/batch |  |
| MFLOPS<sup>c</sup> | 253.652 | 1,454.363 |  |
| Eval. time<sup>b</sup> | 453 ms/batch | 564 ms/batch |  |
|  | Mean (std) of 6 models |  | Ensemble model |
|  | Validation set | Test set | Test Set |
| Accuracy | 0.813 (0.020) | 0.756 (0.015) | 0.789 |
| Precision | 0.740 (0.056) | 0.693 (0.052) | 0.739 |
| Recall | 0.793 (0.089) | 0.676 (0.081) | 0.693 |
| F<sub>1</sub> score | 0.759 (0.025) | 0.678 (0.019) | 0.715 |
| ROC<sub>AUC</sub> | 0.895 (0.007) | 0.828 (0.005) | 0.852 |
| **CONF 2: Module 1 masks + Module 2 without attention** |  |  |  |
|  | Single Model | Ensemble model |  |
| Parameters | 19,441 | 116,646 |  |
| Memory<sup>a</sup> | 0.406 Gb/batch – 20.08 Mb/batch | 30.02 Mb/batch |  |
| MFLOPS | 241.808 | 1,383.295 |  |
| Eval. time<sup>b</sup> | 442 ms/batch | 526 ms/batch |  |
|  | Mean (std) of 6 models |  | Ensemble model |
|  | Validation set | Test set | Test Set |
| Accuracy | 0.824 (0.017) | 0.768 (0.008) | 0.786 |
| Precision | 0.770 (0.040) | 0.729 (0.007) | 0.756 |
| Recall | 0.768 (0.050) | 0.627 (0.033) | 0.649 |
| F<sub>1</sub> score | 0.767 (0.022) | 0.674 (0.019) | 0.698 |
| ROC<sub>AUC</sub> | 0.895 (0.018) | 0.825 (0.011) | 0.849 |
| **CONF 3: Module 1 masks + Module 2 with WST block replaced by 1 CONV RES block** |  |  |  |
|  | Single Model | Ensemble model |  |
| Parameters | 22,753 | 136,518 |  |
| MFLOPS | 263.284 | 1,578.636 |  |
| Memory<sup>a</sup> | 0.537 Gb/batch – 2.76 Mb/batch | 11.83 Mb/batch |  |
| Eval. time<sup>b</sup> | 164 ms/batch | 273 ms/batch |  |
|  | Mean (std) of 6 models |  | Ensemble model |
|  | Validation set | Test set | Test Set |
| Accuracy | 0.770 (0.027) | 0.733 (0.015) | 0.763 |
| Precision | 0.670 (0.046) | 0.651 (0.023) | 0.696 |
| Recall | 0.762 (0.049) | 0.651 (0.022) | 0.675 |
| F<sub>1</sub> score | 0.711 (0.040) | 0.651 (0.017) | 0.685 |
| ROC<sub>AUC</sub> | 0.847 (0.021) | 0.793 (0.017) | 0.820 |
| **CONF 4: Module 1 masks + Module 2 with WST block replaced by 2 CONV RES blocks** |  |  |  |
|  | Single Model | Ensemble model |  |
| Parameters | 31,423 | 188,538 |  |
| MFLOPS | 365.429 | 2,191.505 |  |
| Memory<sup>a</sup> | 0.711 Gb/batch – 3.26 Mb/batch | 14.41 Mb/batch |  |
| Eval. time<sup>b</sup> | 193 ms/batch | 326 ms/batch |  |
|  | Mean (std) of 6 models |  | Ensemble model |
|  | Validation set | Test set | Test Set |
| Accuracy | 0.746 (0.017) | 0.716 (0.012) | 0.742 |
| Precision | 0.621 (0.036) | 0.608 (0.022) | 0.635 |
| Recall | 0.833 (0.036) | 0.739 (0.045) | 0.768 |
| F<sub>1</sub> score | 0.710 (0.022) | 0.666 (0.010) | 0.695 |
| ROC<sub>AUC</sub> | 0.851 (0.017) | 0.796 (0.017) | 0.822 |
| **CONF 5: Module 2 only (no masks, no attention)** |  |  |  |
|  | Single Model | Ensemble model |  |
| Parameters | 19,441 | 116,646 |  |
| MFLOPS | 241.801 | 1,383.282 |  |
| Memory<sup>a</sup> | 0.406 Gb/batch – 20.08 Mb/batch | 26.02 Mb/batch |  |
| Eval. time<sup>b</sup> | 382 ms/batch | 474 ms/batch |  |
|  | Mean (std) of 6 models |  | Ensemble model |
|  | Validation set | Test set | Test Set |
| Accuracy | 0.782 (0.021) | 0.774 (0.008) | 0.792 |
| Precision | 0.658 (0.050) | 0.665 (0.032) | 0.681 |
| Recall | 0.888 (0.055) | 0.837 (0.058) | 0.860 |
| F<sub>1</sub> score | 0.753 (0.016) | 0.739 (0.008) | 0.760 |
| ROC<sub>AUC</sub> | 0.897 (0.009) | 0.875 (0.090) | 0.895 |

In all cases, individual models were trained with 6-fold cross-validation with the HFHS training set of 2265 CXRs. Several hyperparameters (see Supplementary Information Table S1) were fine-tuned to achieve optimal convergence and classification performance of the entire pipeline in its default configuration (Table 1, CONF 1). The same hyperparameters were then used for the training of all alternative configurations of the network shown in Table 1. Each epoch processed 145 batches of 13 training images and 29 batches of 13 validation images. The augmentation strategy was similar to that already described for the training of Module 1, with the notable difference that both training and validation images, and their corresponding masks (if used) calculated by Module 1, were augmented, and thus, in each epoch, the network trained on 1885 (145 × 13) different augmented images/masks, and was validated against 377 (29 × 13) different augmented images/masks. Based on the trend of the validation loss and accuracy, training was stopped after 200 epochs in each validation run. Individual models derived from each run were then combined into an *ensemble* model (Supplementary Information Fig. S8) without averaging their layers coefficients. In our default pipeline (Table 1, CONF 1) the final ensemble model contained a single WST block followed by 6 parallel branches, each with 1 ATTENTION and 3 CONV RES blocks. The random selection of the training and validation images in each run is shown in Fig. 8a. The training and validation loss and accuracy for each run are shown in Fig. 8b. Receiver-Operating Characteristic (ROC) curves for the validation images in each run are shown in Fig. 8c; a confusion matrix combining all validation images is shown in Fig. 8d. The Covid vs. non-Covid classification performance of the ensemble model with the non-augmented test sets of 1532 CXRs is displayed in terms of ROC curves and Confusion Matrices in Figs. 8e,f.

**Fig. 8.**
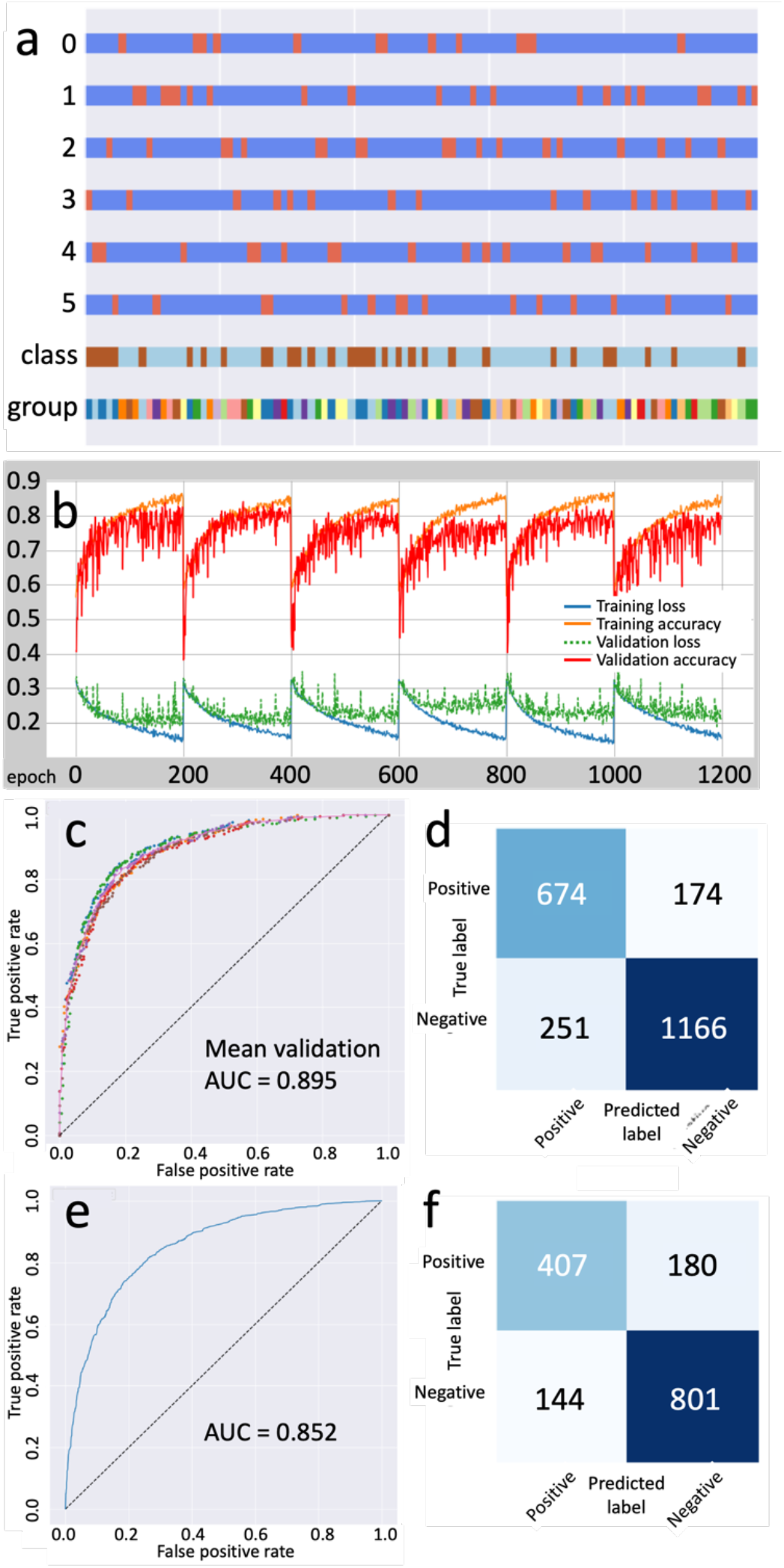
CXR-Net training (default configuration, Table 1, CONF 1). *Panel a*: partition of 2265 CXRs into 6 randomly selected train (blue) and validation (orange) subsets (rows ‘0-5’). Cyan and brown sections (row labeled ‘class’) display the Covid +/– partition. A multicolor row (row labeled ‘group’) displays the partition between patients. *Panel b*: loss and categorical accuracy in the 6 training and validation sets. *Panel c*: validation ROC curves for each run (circles), and mean validation ROC curve for all 6 runs (continuous line). *Panel d*: confusion matrix for the combined validation sets. *Panels e,f*: ROC curve and confusion matrix for the ensemble model predictions with non-augmented 1532 CXRs of the test set.

One-way analysis of variance (Anova) (Fig. 9, Supplementary Information Table S2) of the 6 cross-validated training runs with different Module 2 configurations, reveals that the ATTENTION block does not contribute significantly to the F1 score or the ROC area under the curve (auc), although some small positive effect can still be appreciated in the performance of the ensemble model (Table 1, compare CONF 1 and 2). For this reason, until further tests with outside data clarify whether this block is useful or not, we retain it in the default configuration of the network (Table 1, CONF 1). Instead, statistically significant differences are noted between the default configuration and the configurations in which either the WST block is removed and replaced by 1 or 2 CONV RES blocks (Table 1, CONF 3 and 4), or lung masks are not used (Table 1, CONF 5). In particular, removing the WST block degrades the F1 and ROC values, as we would expect from the loss in image generalization akin to transfer learning provided by the WST. This loss is insufficiently replaced by adding more convolutional layers to the network. In contrast, somewhat unexpectedly, removing the use of lung masks significantly improves the F1 and ROC values. We have further investigated the meaning of the latter result through the use of Grad-CAM saliency (heat) maps [9].

**Fig. 9.**
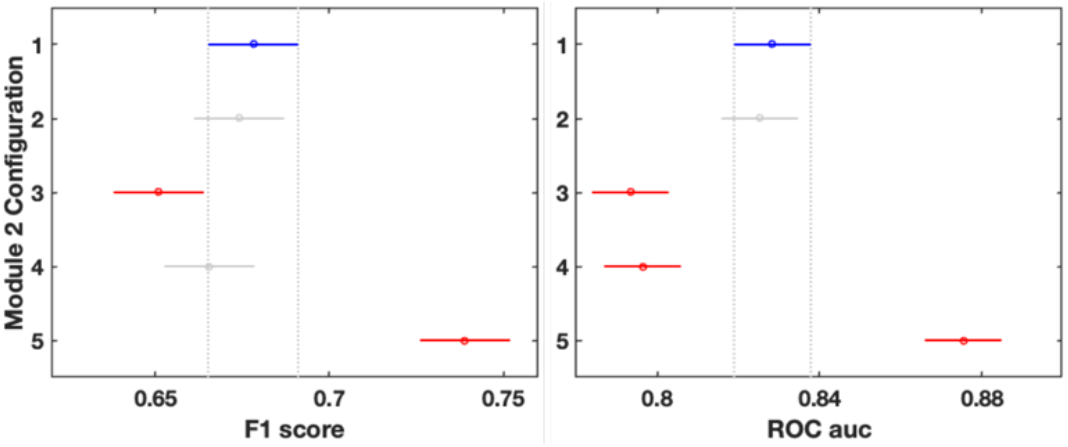
Anova based comparison graph of 6 cross-validated models with different Module 2 configurations (Table 1), using images in the test set. Each group mean and interval are represented by a full circle symbol, and a line extending out from the symbol. The first group mean and interval are highlighted in blue. The intervals for the other groups that do not intersect with the interval for group 1 mean, are highlighted in red. F1 score: 2 groups have means different from groups 1 (p=4.04e-9). ROC auc: 3 groups have means different from groups 1 (p=8.20e-12).

### D. Heat maps

Grad-CAM saliency maps [9] often suffer from low resolution, and thus fail to differentiate multifocal lesions within an image. CXR-Net Module 2 feature maps retain the same spatial dimensions and resolution throughout all layers, including the final convolutional layer that is used to generate the saliency maps. The two channels of this layer contain the predictions for the CXR image probability of being Covid or non-Covid, respectively, summing to 1. We generate Grad-CAM maps from both channels. When Grad-CAM maps were generated by Module 2 in the configuration that uses lung masks (Table 1, CONF 1), texture alterations in the lung regions of the CXRs (leftmost column in Fig. 10) were readily identified as the ‘hot’ regions of the saliency maps (middle column in Fig. 10). In contrast, when Grad-CAM maps were generated by Module 2 in the configuration that does not use masks (Table 1, CONF 5), despite the apparently superior classification performance of this configuration, the hot regions of the maps were mostly located outside the lung regions (i.e., diaphragm, shoulder, labels, image edges). This observation is consistent with similar observations in [13, 14, 16], suggesting that in the absence of information from lung segmentation the network relies heavily on confounding features that originate during the CXR acquisition, or that systematically differ between Covid and non-Covid datasets. This interpretation is confirmed by an analysis of the distribution of the class probabilities predicted with the test set when Module 2 operates with or without lung masks (Fig. 11). In its default configuration (Table 1, CONF 1) the ensemble model of CXR-Net predicts significantly better non-Covid than Covid CXRs (Fig. 11a). In the absence of information from lung masks, predictions for the Covid class present the highest median and the smallest variability of all configurations, while the median of the predictions for the non-Covid class is decreased and its variability increased (Fig. 11b). This behavior is exactly what would be expected if the Covid class is confounded by systematic ‘Covid’ signals (markings, age, sex, race, position, etc.) in the non-lung regions of CXRs, while the non-Covid class, being simply assembled from a variety of pre-Covid era CXRs, is heterogenous with respect to non-lung regions. The ‘outliers’ distributions also supports this interpretation. In fact, when only the lung regions are considered (Fig. 11a), many CXRs from pre-Covid era that contains features of pneumonia similar to SARS-CoV-2 are predicted as non-Covid with very low probability. When both lung and non-lung regions are considered (Fig. 11b) the contribution of pneumonia features in non-Covid CXRs is diluted in the set, lowering the median and decreasing the number of outliers. Covid patients whose non-lung regions do not contain common systematic features appears as outliers.

**Fig. 10.**
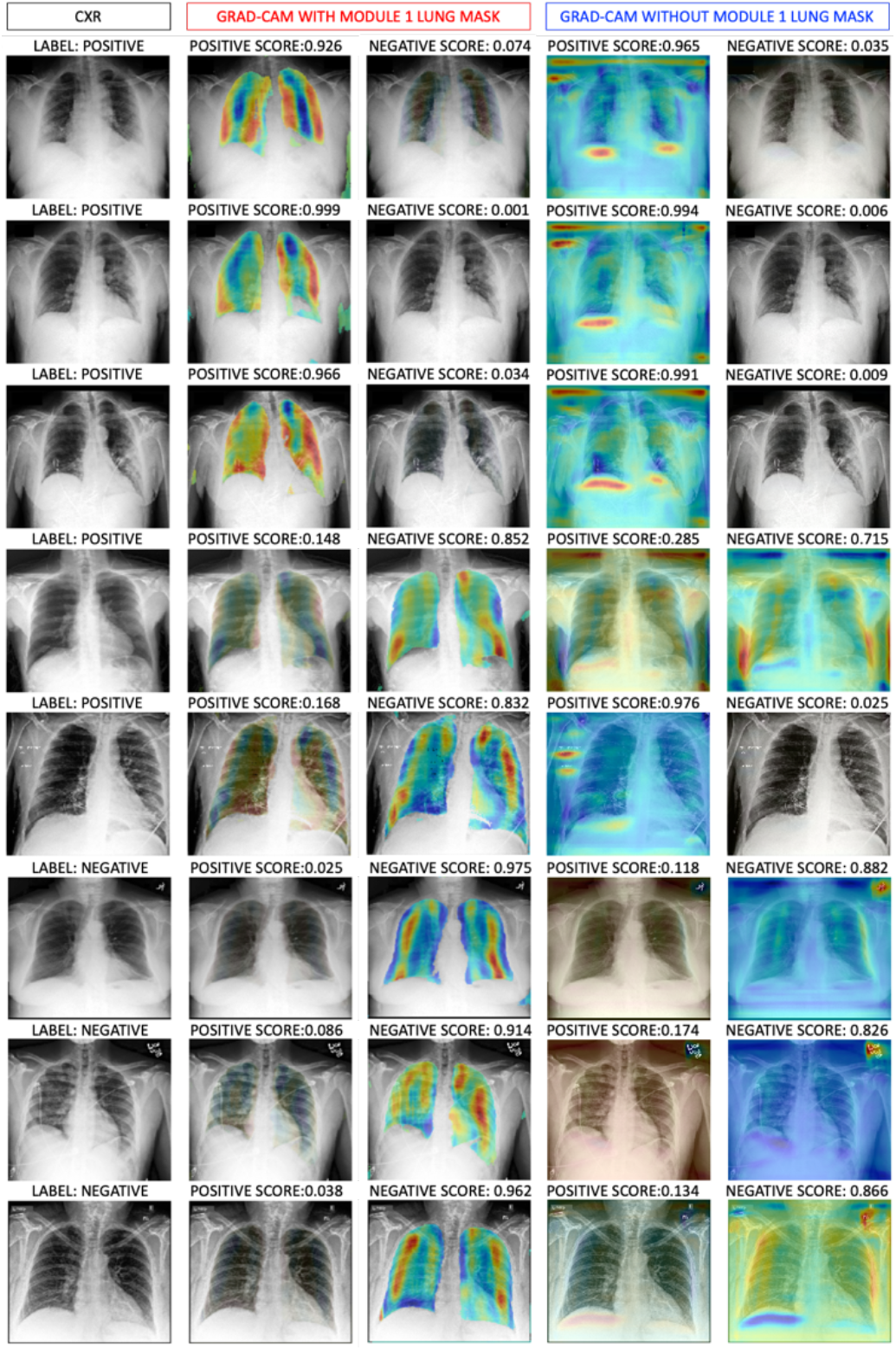
Ensemble model predictions for some CXRs in the HFHS test set. The leftmost column shows the input CXRs and labels (positive for Covid and negative for non-Covid) based on image analysis and RT-PCR. The 2^nd^ and 3^rd^ columns from the left show the model assigned positive and negative scores, and the corresponding Grad-CAM maps calculated using lung masks (Table 1, CONF 1). The 4^th^ and 5^th^ columns from the left show the model assigned positive and negative scores, and the corresponding Grad-CAM maps calculated without using lung masks (Table 1, CONF 5). The top 3 rows shows a correct positive assignment by both models. The 4th row shows a CXR labeled positive, but assigned as negative by both models. The 5th row shows a CXRs labeled positive, but incorrectly assigned as negative when the lung masks are used. The bottom 3 rows shows CXRs labeled negative, and similarly assigned by both models.

**Fig. 11.**
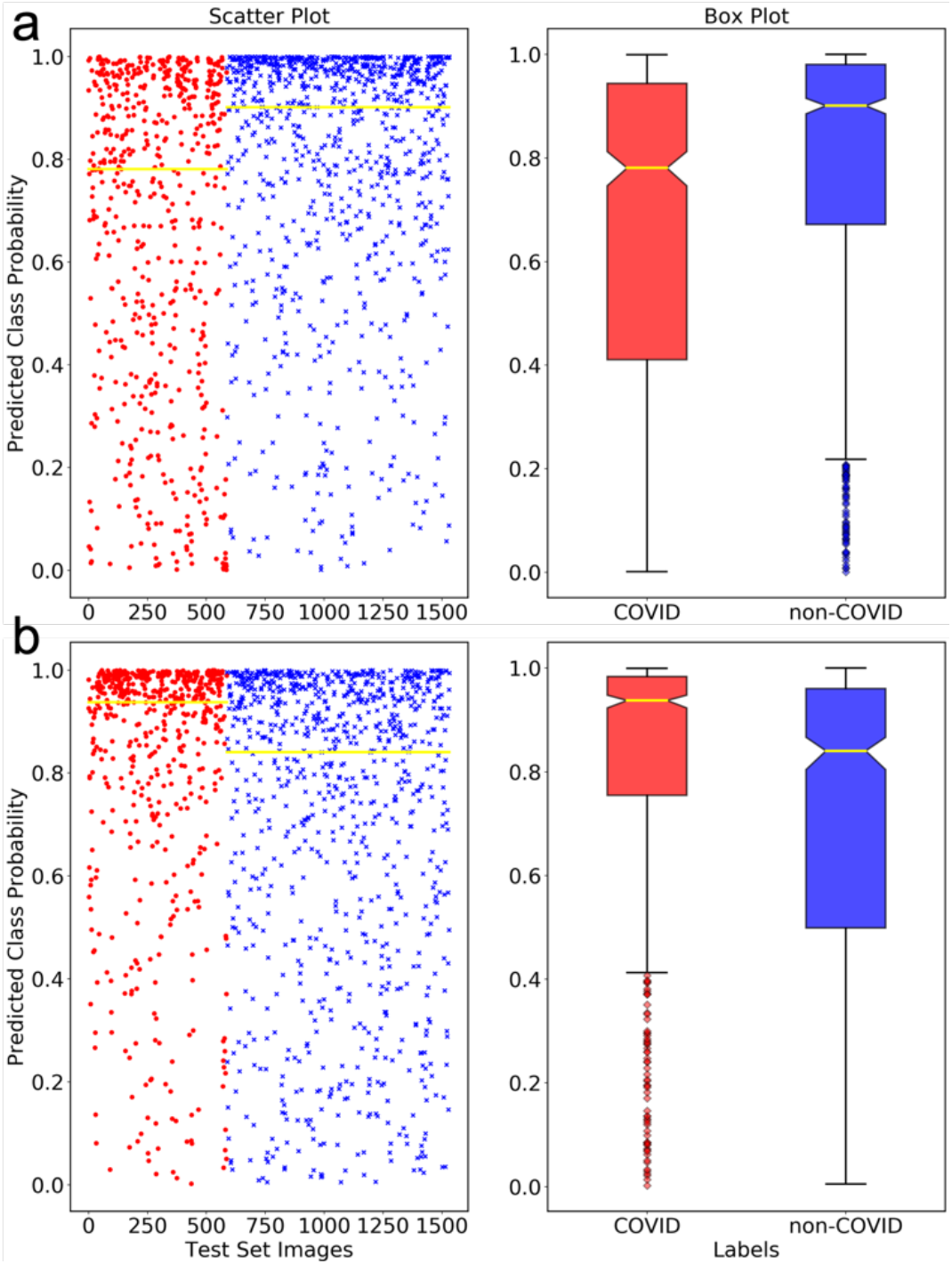
Anova based scatter and box plots of the distributions of Covid *vs* non-Covid prediction probabilities assigned by the ensemble model of CXR-Net in different configurations. a. Module 1 masks + Module 2 (Table 1, CONF 1). b. Module 2 only (Table 1, CONF 5). Red dots and boxes show the predicted probabilities of being Covid for CXRs that are truly Covid. Blue dots and boxes show the predicted probabilities of being non-Covid for CXRs that are truly non-Covid. Medians of the distributions are shown as yellow lines.

### E. Graphic interphase

We have developed g-CXR-Net [41], a graphic front-hand for CXR-Net ensemble model, which adopts the Python *<u>tkinter</u>* package as interface to the Tk GUI toolkit (https://www.tcl.tk/), and is compatible with most personal computer platforms (Fig. 12). CXR images of any size in *dicom* format are classified as *Covid* or non-*Covid*, and lung heat maps are generated in seconds on a desktop, laptop, or hand-held device.

**Fig. 12.**
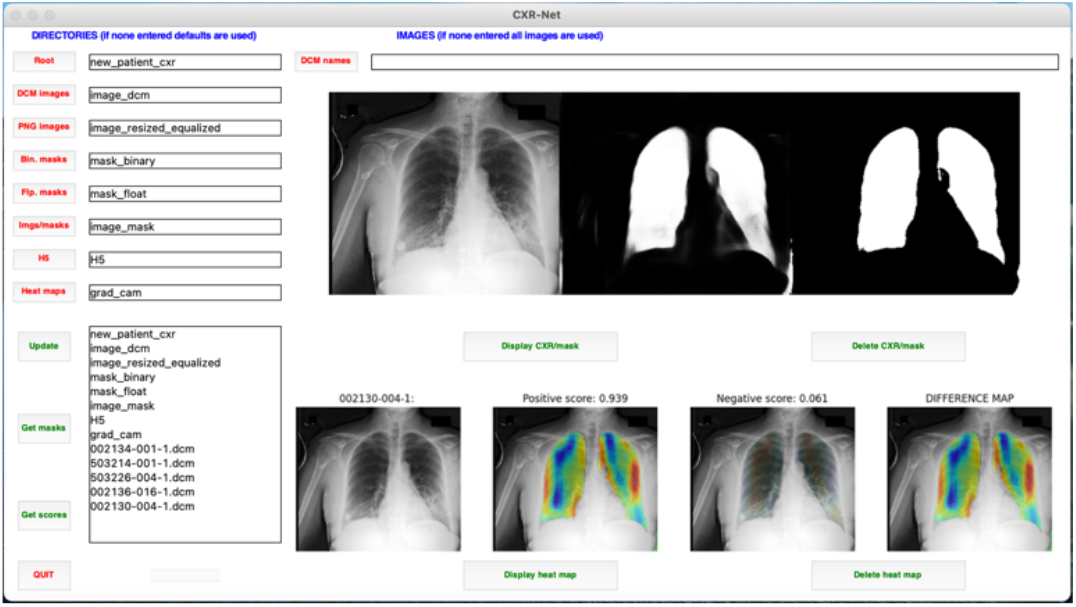
g-CXR-Net GUI. Buttons are associated with different actions: a) select a directory tree for sources and targets different from the default one, b) select CXR images to be processed, c) update selections, d) calculate lung masks, e) calculate classification scores, f) calculate heat maps, g) display CXRs and corresponding floating point and binary lung masks, h) display heat maps, i) quit. Updates and task results are displayed in the text window at the lower left corner of the GUI.

## IV. Limitations of the study

CXR-Net features a dual-module architecture for the sequential segmentation of the lung fields in AP/PA CXRs, and their classification as either non-Covid (normal or other non-Covid pathology) or SARS-CoV-2. We have shown that lung segmentation is necessary to avoid an artifactual identification of Covid cases on the basis of systematic features of non-lung regions that may be common in CXRs of Covid patients. For this reason, despite the higher classification performance of Configuration 5 (Table 1, CONF 5), we recommend Configuration 1 as the preferred architecture. However, it should be noted that the lung segmentations provided by Module 1 are still very inaccurate when a pathologic consolidation of the lung tissue produces areas with pixel intensity comparable to those of non-lung tissues. In extreme cases, an entire affected lung can be excluded from the mask of both lungs, leaving in the mask only the unaffected lung, and thus leading to an incorrect classification. This is a serious problem, for which a variety of solutions have been proposed. Carvalho Souza *et al*. [42] have used a first NN to derive an initial segmentation that excludes lung opacities, and a second NN to reconstruct the lung regions “lost” due to pulmonary abnormalities. Selvan *et al*. [43] have treated high opacity regions as missing data and designed an image segmentation network that utilizes a generative model based on a variational encoder for data imputation. Motamed *et al*. [20] and Munawar *et al*. [15] have used generative adversarial networks (GANs) for both lung segmentation and classification. In this case, the GAN generator is trained to generate a segmented mask of a given input CXR. The discriminator distinguishes between a ground truth and the generated mask, and updates the generator through the adversarial loss measure. Despite these efforts, when patients’ pathologies lower the contrast between lungs and surroundings in the CXR, the segmentation task remains very challenging, and a generally accepted strategy to improve the outcome has not been identified yet.

Module 2 is a hybrid convnet that uses the Wavelet Scattering Transform (WST) of the input image [24, 25, 34] as the first layer. Since the coefficient of the WST block are not learned, Module 2 requires the refinement of only ∼21,000 parameters for images of dimensions 300 × 340. For this reason, in this study we could combine without parameter averaging the models derived from 6 cross-validated runs into a single ensemble model (Supplementary Information Fig. S8) of only ∼123,000 parameters. In our experience, retaining the individual Module 2 validation models as parallel paths, rather than averaging their refined coefficients, improves the final classification performance of the ensemble model (Supplementary Information Table S1). Currently, the kernel sizes of the convolution operations inside the CONV RES blocks of Module 2 are optimized for the feature maps produced by the WST block. We have found that using CXR input images larger than 300 × 340 only slows down training without improving classification. However, were a user to opt for larger images, a manual optimization of the kernels inside the CONV RES block would be required. Future releases of the pipeline will adapt automatically the kernel sizes to the dimensions of the input images.

The subset of non-Covid CXRs used in this study was selected out of the HFHS archive of images from pre-Covid era, and contains images of normal lungs as well as lungs affected by non-Covid pathologies, some of which, however, resemble Covid pneumonia. Since the current model was trained only on the Covid and non-Covid subset of images, it allows for a fast identification of Covid cases based on a simple binary choice. However, this type of training is also expected to increase the number of *false negative* attributions. Furthermore, while the pipeline can be easily modified for CXR classification into normal and various lung pathologies by increasing the number of channels of the last convolutional layer of Module 2, we can expect that, as classes multiply, the performance of the pretrained model will drop [44]. This is an issue of general importance in automated Covid detection from CXRs, because Covid progression entails at least three distinct stages (early infection, pulmonary phase, and hyper-inflammation phase) with distinct radiological features [45-49]. Unfortunately, while the HFHS database contains information on the time difference between the date of confirmed Covid diagnosis and the date of CXR acquisition, it does not contain annotations about the disease stage, or alteration in specific lung zones that would support the stage assignment. As a consequence, CXRs acquired at some late hospitalization time, which may contain features of recovery from Covid, are labeled the same way as others acquired at an earlier time, showing severe pulmonary involvement. Additional work will be required to include in the HFHS database appropriate metadata that report the exact stage of the disease in addition to the mere diagnosis.

A different problem in the HFHS database is represented by the fact that the Covid subset is still significantly smaller than the non-Covid subset. To relieve this problem, during training CXR-Net uses traditional techniques of data augmentation based on image rotation, sheering, shifting, scaling, and mirroring. However, it has been reported that synthetic normal and Covid CXRs produced by GANs can be utilized to enhance further the performance of Covid detection [50, 51], and these methods will be implemented in future releases of the pipeline.

## Data Availability

All data produced in the present work are contained in the manuscript

