## Supplementary Information for "A Hybrid Pipeline for Covid-19 Screening Incorporating Lungs Segmentation and Wavelet Based Preprocessing of Chest X-Rays"

**Figure S1. Module 1 architecture**

Six residual CONV RES blocks are repeated in a linear path along which the dimensions of the feature maps remain identical to those of the input image and of the output mask(s). The residual path of a CONV RES block consists of three parallel branches of separable/atrous convolutions. Parallel branches inside the residual block are concatenated before adding them to the shortcut connection. A Spatial Dropout layer follows each residual block. In this study we have used 6 CONV RES blocks, each with kernel sizes of [3,3], [5,5], [7,7], and dilation rates of [1,1], [3,3], [5,5], respectively, with 16 filters in each residual branch, and 48 filters in the shortcut branch.

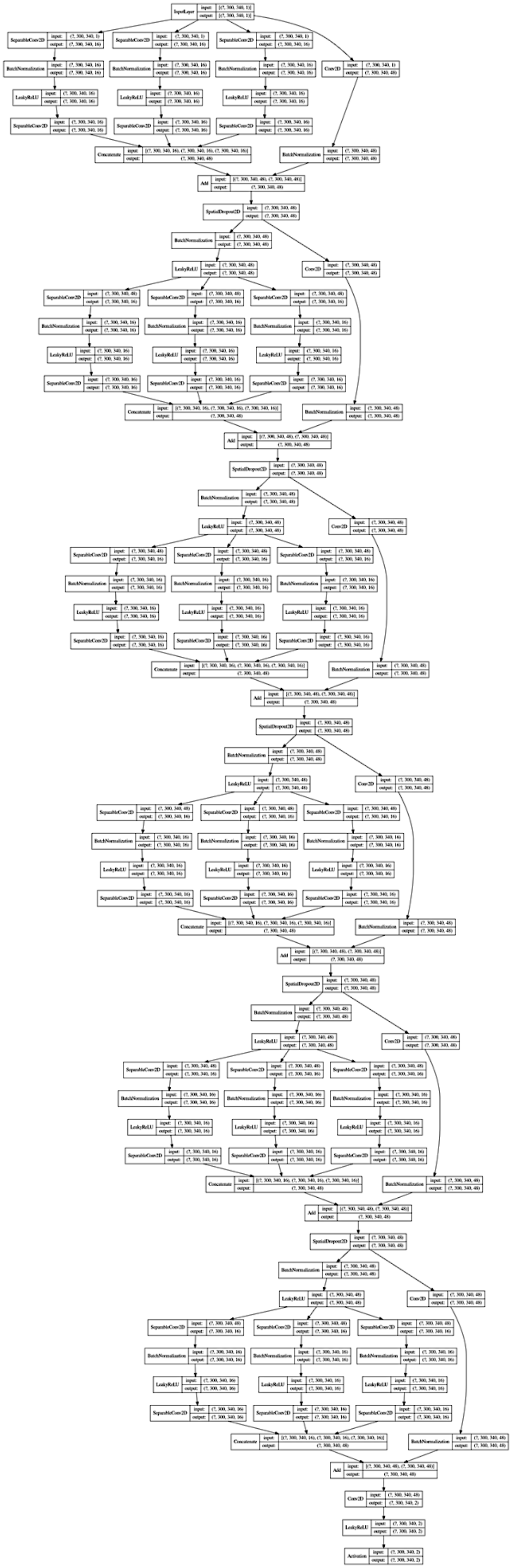

**Figure S2. Calculation of cross-attention between the input CXR and the lung masks.**

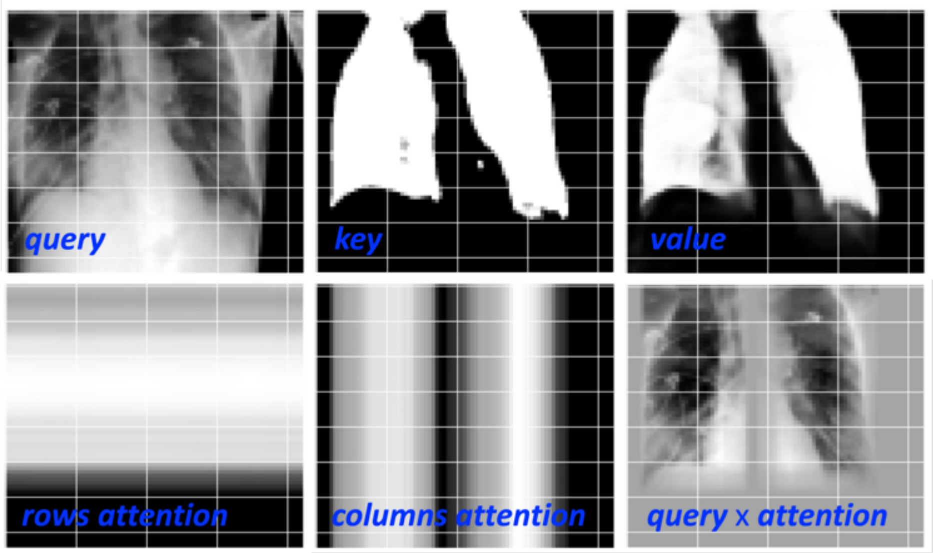


The down-sampled CXR (top left), lungs binary mask (top middle), and lungs floating point mask (top right), are the *query*, *key*, and *value* features, respectively, used to calculate the cross-attention along the image rows (bottom left) between the CXR and the lung regions in a Multi-head Attention layer (2 heads of size 64). Cross attention along the image columns (bottom middle) is calculates using the *query*, *key*, and *value* transposes. Rows and columns cross attentions are multiplied pointwise by the *query* image to obtain the row- and column-wise CXR attention (bottom right).

**Figure S3. Module 2 architecture**
**Configuration 1**

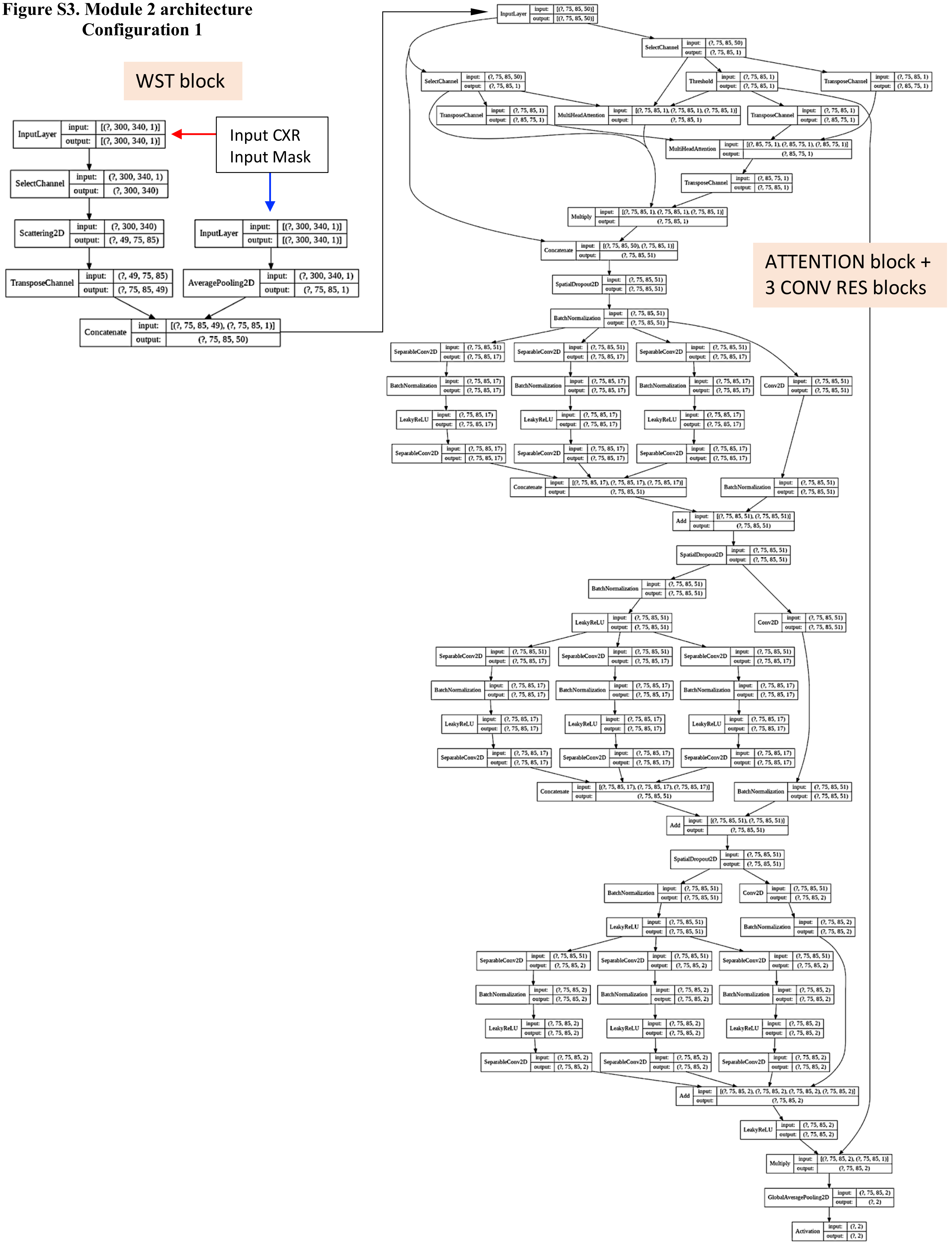

**Figure S4. Module 2 architecture**
**Configuration 2**

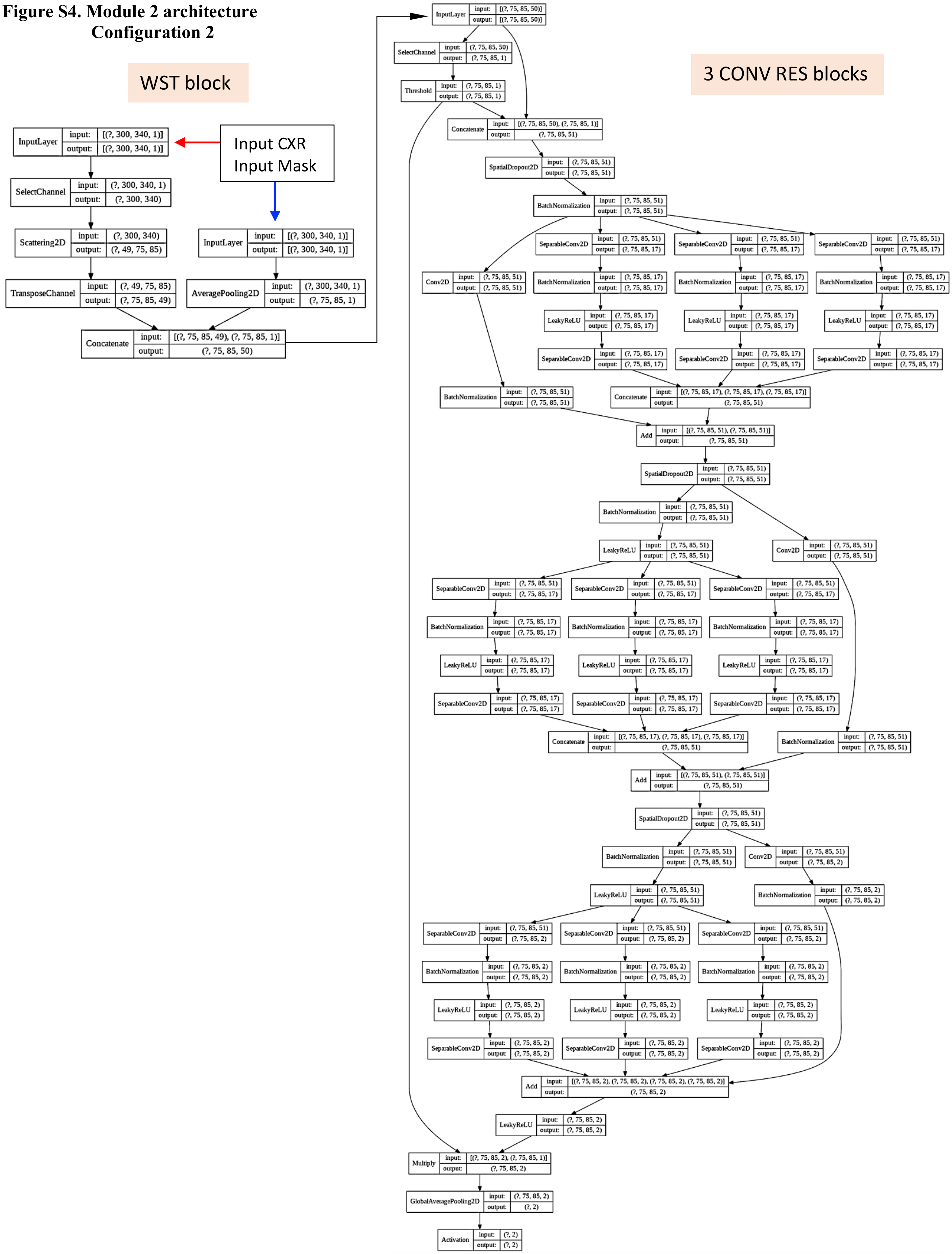

**Figure S5. Module 2 architecture**
**Configuration 3**

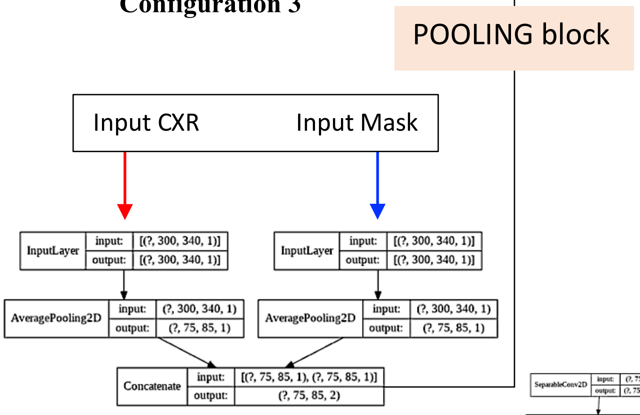


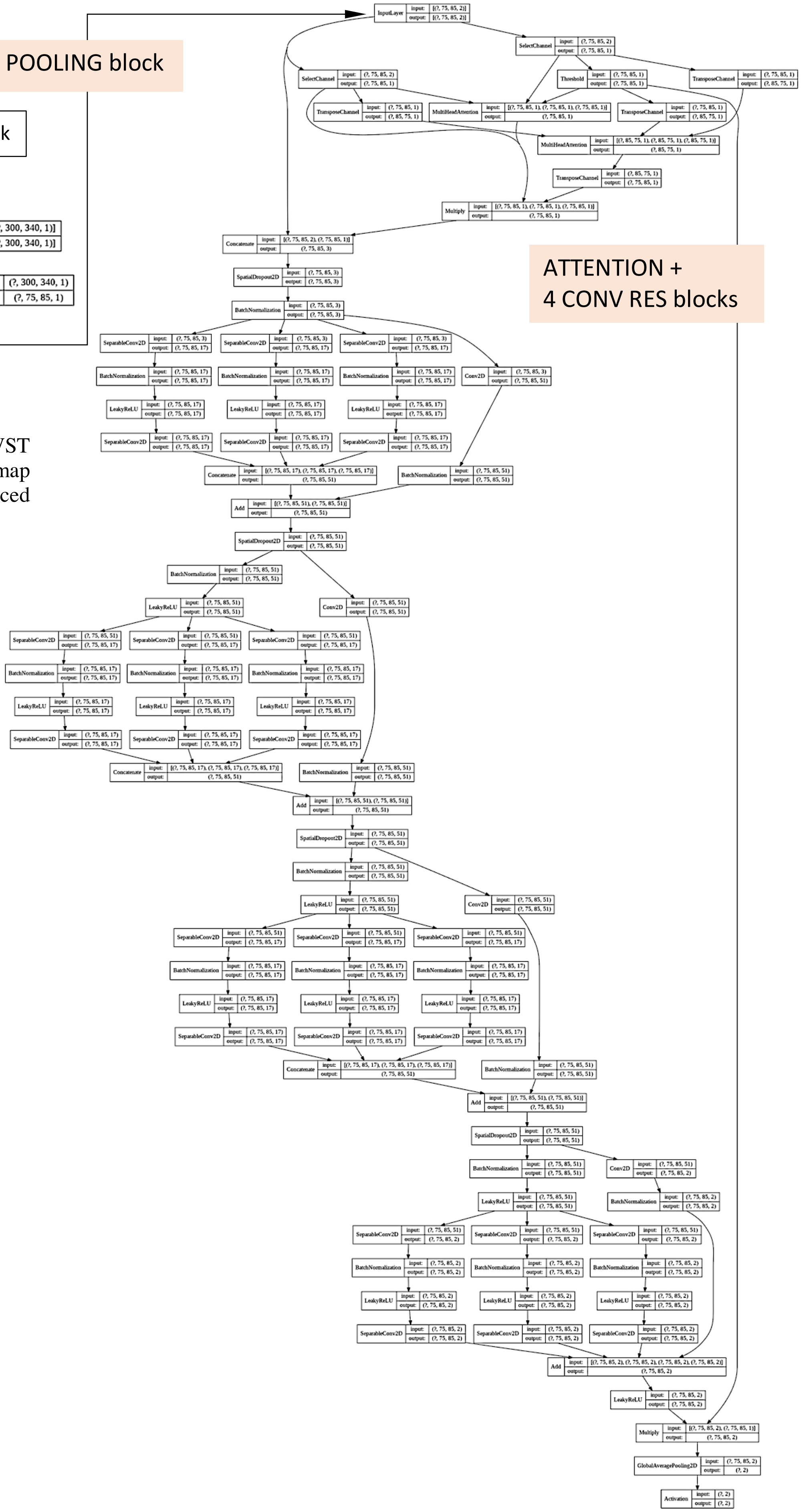


A POOLING block replaces the WST block in order to produce a feature map of the same dimensions as that produced by the WST block.

**Fig. S6. Module 2 architecture**
**Configuration 4**

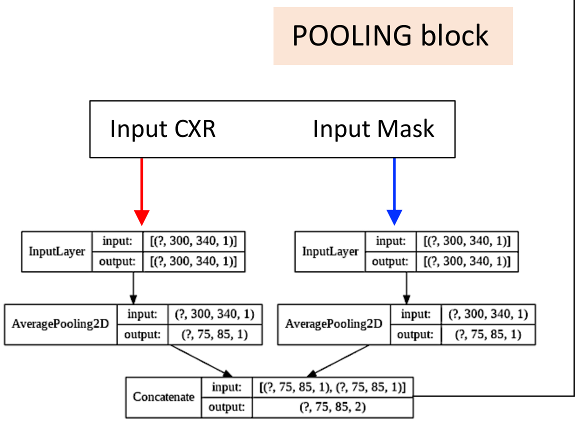


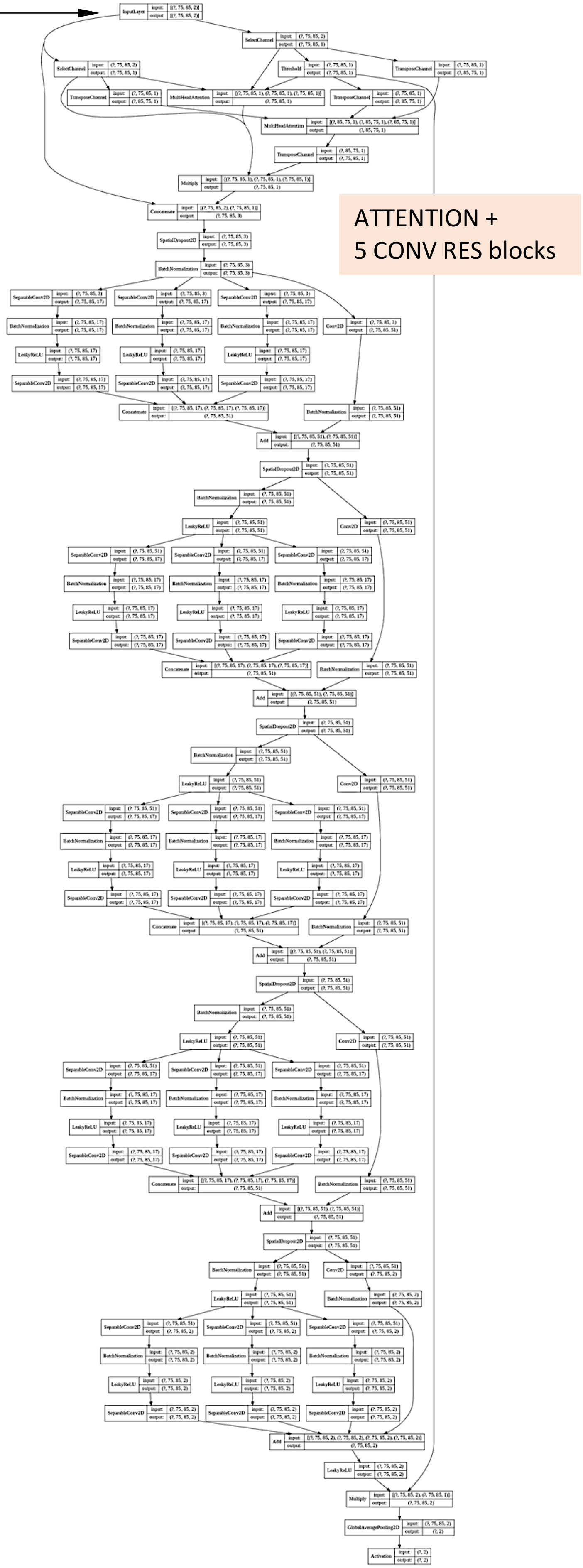


A POOLING block replaces the WST block in order to produce a feature map of the same dimensions as that produced by the WST block.

**Fig. S7. Module 2 architecture**
**Configuration 5**

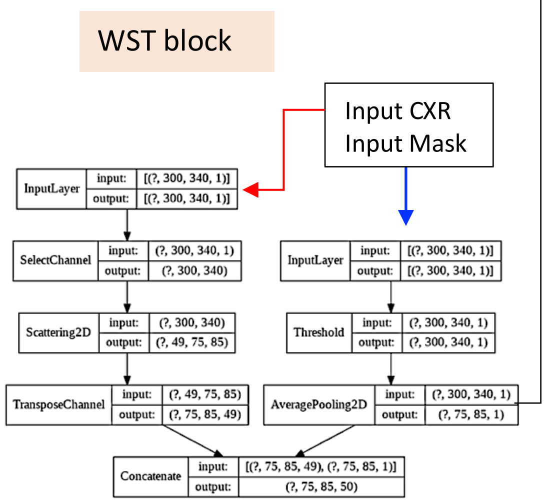


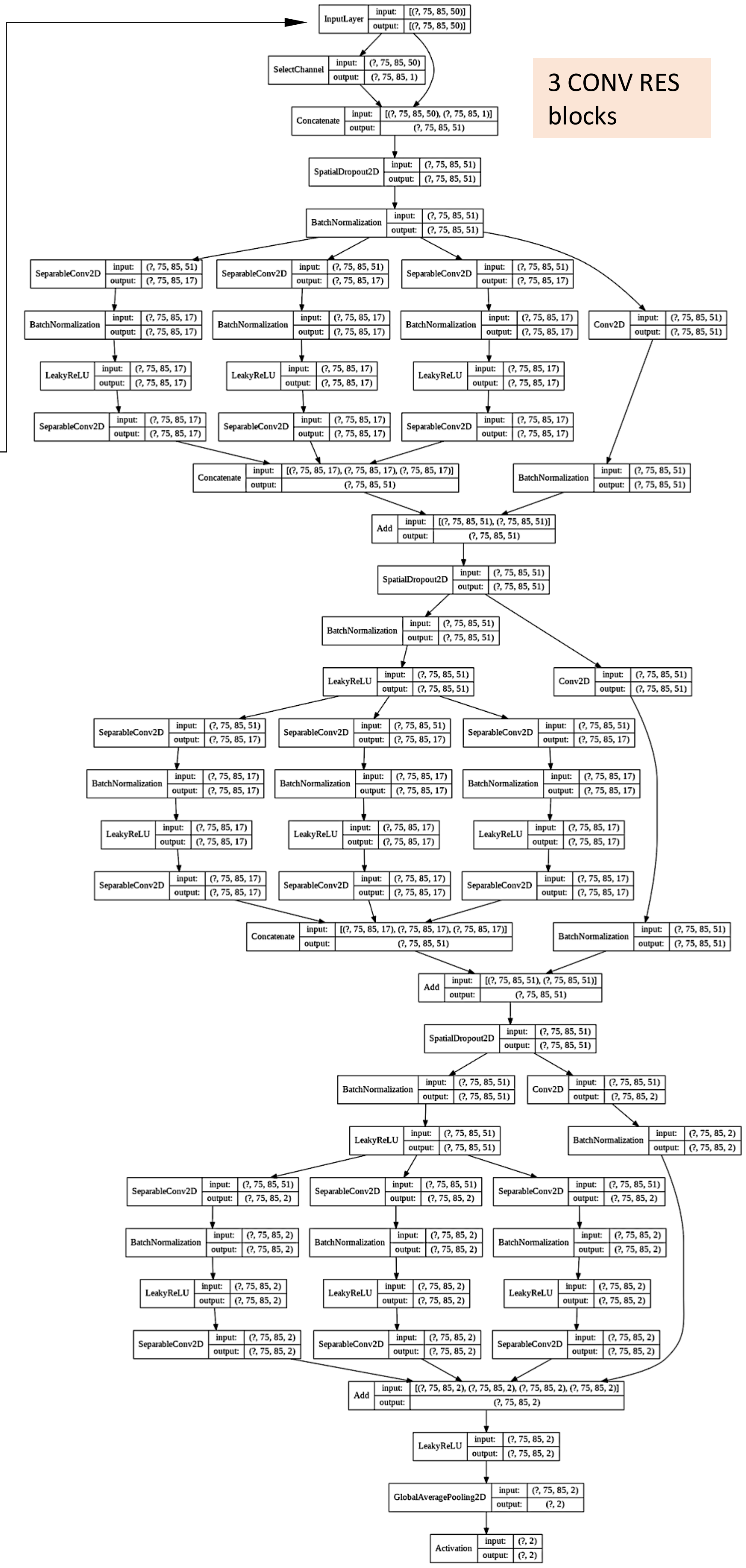


In order to retain the same spatial dimensions of the output from the original WST block, a Thresholding operation converts the input lung mask to a feature map with all pixels equal to 1. This operation effectively removes the mask information from training and from forward pass validation.

**Figure S8. Ensemble model architecture**

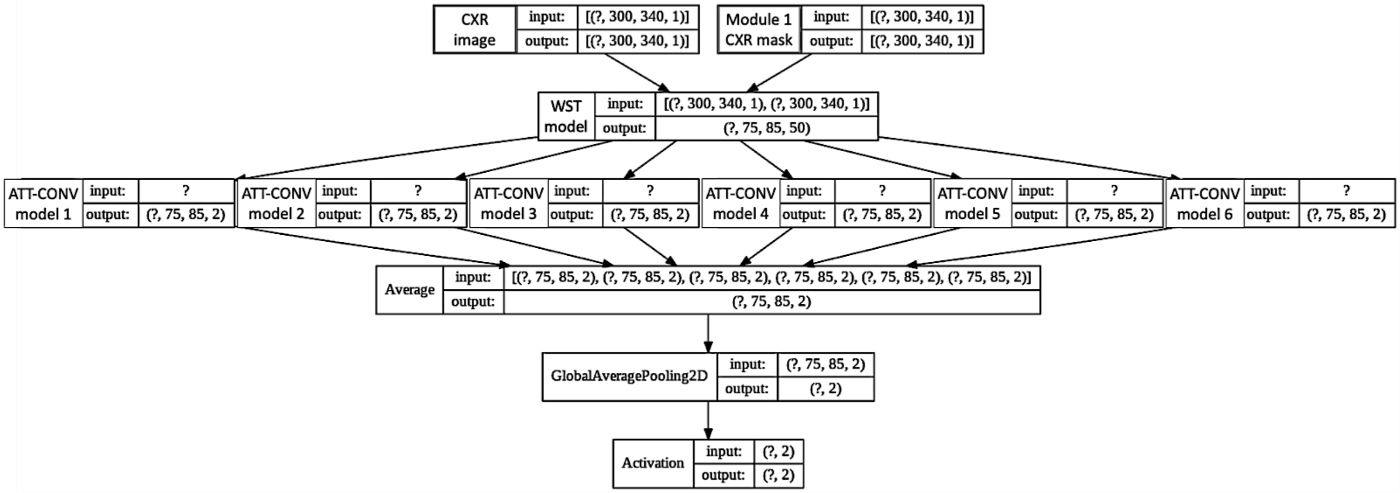


A single WST block inputs 6 parallel ATTENTION + CONV RES blocks, whose output is averaged to a single feature map of dimensions $75 \times 85 \times 2$. A GlobalAveragePooling2D layer passes this map to the final Softmax activation layer. Question marks represent the number of images in each batch.

**Table S1.**

TABLE S1. MODULE 2 HYPERPARAMETERS SEARCH

| Parameter | Range Explored | Optimized value |
|---|---|---|
| WST block | | |
| J | 2-3 | 2 |
| L | 4-8 | 6 |
| ATTENTION block | | |
| No. of heads | 1-5 | 2 |
| Query, key, and value vector size | 32-96 | 64 |
| CONV RES blocks | | |
| No. of blocks | 2 to 7 | 3 |
| No. of filters | 12 to 24 | 17 |
| Size of each of 3 Kernels in the residual path (Fig. 6) | (3,3) - (7,7) | (3,3),(3,3),(3,3) |
| Dilation of each of 3 Kernels in the residual path (Fig. 6) | (1,1) - (5,5) | (1,1),(2,2),(3,3) |
| Drop out rates | 0.05 – 0.1 | 0.1 |
| OPTIMIZER | | |
| Method | Adam | Adam |
| Learning rate | 0.0005-0.01 | 0.002 |
| Beta1 | 0.7-1.0 | 0.899 |
| Beta2 | 0.7-1.0 | 0.999 |
| Epsilon | 1e-9 -1e-5 | 1e-7 |
| Amsgrad option | False-True | True |
| ENSEMBLE GENERATION | | |
| Coefficients Averaging | False-True | False |

**Table S2**

TABLE S2. *P* VALUES FROM THE ANOVA COMPARISON OF 6 CROSS-VALIDATED CXR-NET INDIVIDUAL MODELS WITH THE TEST SET

| Group | **1** | **2** | **3** | **4** | **5** |
|---|---|---|---|---|---|
| **1** | | 0.9878 | 0.0001 | 0.0004 | 0.0000 |
| **2** | 0.9903 | | 0.0004 | 0.0012 | 0.0000 |
| **3** | 0.0363 | 0.0964 | | 0.9896 | 0.0000 |
| **4** | 0.6162 | 0.8666 | 0.4788 | | 0.0000 |
| **5** | 0.0000 | 0.0000 | 0.0000 | 0.0000 | |

The matrix contains the *p*-values for the hypothesis test that the corresponding differences between the group means (as shown in Figure 9) are equal to zero. Values below and above the diagonal refer to the F1 score and ROC_auc values, respectively.
